# Influence of Binarization Process on Vascular Density Metrics: A Quantitative Optical Coherence Tomography Angiography Assessment in Human and Porcine Retinas

**DOI:** 10.64898/2025.12.11.25342086

**Authors:** Irtiza Sakif Islam, Jai Singh Rajput, Kasandra Albarran, Syed Faaiz Enam, Mahabad Barwari, Aksharkumar Dobariya, Ankur Patel, Misha Dunbar, Juan Pascual, Ulrike Hoffmann, Sourav S. Patnaik

**Affiliations:** Department of Anesthesiology and Pain Management, The University of Texas Southwestern Medical Center, Dallas, TX, USA; Department of Neurology, The University of Texas Southwestern Medical Center, Dallas, TX, USA; Department of Physiology, Biophysics & Systems Biology, Weill Cornell Medicine, Cornell University, New York, NY, USA; Department of Neurological Surgery, The University of Texas Southwestern Medical Center, Dallas, TX, USA; Animal Resource Center, The University of Texas Southwestern Medical Center, Dallas, TX, USA; Division of Child Neurology, Weill Cornell Medicine, Cornell University, New York, NY, USA

**Keywords:** Optical Coherence Tomography Angiography, Vascular Density, Binarization Algorithms, Image Processing, Global Thresholding, Local Adaptive Thresholding, Superficial Capillary Plexus, Deep Capillary Plexus

## Abstract

**Background:** Optical Coherence Tomography Angiography (OCTA) provides high-resolution visualization of retinal microvasculature, with vascular density (VD) serving as a one of the key quantitative metrics. However, VD measurements are highly sensitive to image binarization step, and no standardized approach exists.

**Methods:** We analyzed 51 OCTA scans (human and porcine) using 29 binarization algorithms, including global and local thresholding techniques from ImageJ and DoxaPy, as well as Random Walker segmentation. VD was calculated for each binarization algorithm and compared against Optovue-generated values (ground truth). Results were evaluated using hierarchical clustering and agreement between them was determined by Bland–Altman analysis.

**Results:** Wolf algorithm was found to exhibit least deviation from mean Optovue VD values for human SCP layer (46.5 ± 1.2% vs. 48.3 ± 1.4%; *p* = 3.62 x 10^-5^); however, there is not significant difference between VDs from Optovue and Wolf algorithms from porcine SCP layer (46.2 ± 1.8 % vs 46.3 ± 1.4% ; *p*=0.74). For DCP layer, Phansalkar algorithm exhibited least VD variability (50.7 ± 2.0% vs. 51.9 ± 1.7%; *p*=2.53 x 10^-4^) in the human cohort. Whereas Percentile algorithm exhibited least, non-significant variations in the porcine DCP layer VD (50.0 ± 1.4 % vs. 50.3 ± 1.4%; *p*=0.75).

**Discussion:** Each binarization technique evaluated in this study impacts OCTA-derived VD measurements differently. Local adaptive algorithms collectively outperform global methods, particularly for SCP analysis. Standardization of image processing pipelines and layer-specific optimization are essential to improve reproducibility and clinical consistency.

## Introduction

Optical Coherence Tomography Angiography (OCTA) is a novel non-invasive, dye-less approach of visualizing retinal blood flow and it has exhibited clinically lucrative translation potential as a diagnostic tool for pathologies other than ocular [1–3]. Retinal vascular density obtained via OCTA imaging has been acknowledged as a “reflection” of underlying pathological and microvascular changes in the brain as it shares vasculature with intracranial circulation [3–6]. Further, visualization of cerebral blood flow is difficult whereas retinal blood flow assessment is convenient, and vascular perfusion status serves as a useful biomarker for monitoring critical pathophysiological conditions [3, 4, 6].

Perfusion status of the retinal tissue through OCTA imaging technique is semi-quantitatively assessed via several biomarkers such as vascular density (VD) or blood vessel density, foveal avascular zone area, vessel length density, fractal dimensions, etc. [7, 8]; however, there is no standardized protocol or harmonized processing method that is widely accepted for OCTA biomarker quantification. Vascular density (VD), which is the proportion of the perfused vasculature area to the total scanned retinal tissue area, serves as a an effective surrogate biomarkers of retinal perfusion status [9, 10]. Despite the clinical relevance of VD (and other OCTA biomarkers), its calculation is highly sensitive to the image processing pipeline, particularly the binarization method applied to *en face* OCTA images. Binarization involves converting grayscale images into binary representations to delineate vessels from background tissue. The choice of binarization algorithm can significantly affect the apparent vessel structure, leading to the greatest variability in computed VD values, vessel diameters, and other derived measurements. This is further compounded by the lack of standardization across studies and devices as there is no single thresholding technique that is considered gold standard for OCTA [11, 12]. Furthermore, the information regarding exact steps utilized in image processing (including binarization) to obtain VD values from commercial softwares, is not readily available. Thereby, limiting reproducibility and cross-study comparisons across OCTA laden investigations.

Given the variability in biomarker quantification, device-based variability, and other issues in OCTA imaging [10, 11, 13–22], it is critical to understand the fundamental role played by the binarization step in biomarker quantification. Developments in image processing tools such as ImageJ and Python offer a broad array of binarization strategies, including both global and local (adaptive) thresholding algorithms. Yet, there no single method that is fit for all the retinal layers or experimental models universally. Hence, it is imperative that we understand the effect of binarization techniques on OCTA-derived vascular density metrics in human and porcine retinal scans. For this study, we investigated the performance of 29 different binarization algorithms which included default ImageJ binarization, a Random Walker-based method, and multiple algorithms available in the open-source DoxaPy package and ImageJ software (NIH) [23–49]. We assessed the variability of each binarization technique against values obtained from the Optovue system (primary acquisition device), which were considered ground truth. Our study aimed to identify the most reliable binarization technique (global vs. local) for optical coherence tomography angiography image processing. This determination is a critical step toward the standardization and harmonization of OCTA image processing and biomarker quantification protocols.

## Methods

Detailed experimental steps performed for OCTA image processing in this study is shown in Fig.1. For this study, we utilized an established OCTA protocol for image acquisition from both human and porcine models [6, 50, 51]. We focused on the effect of binarization techniques on the vascular density (VD) metric quantification from the retinal superficial capillary plexus (SCP) and deep capillary plexus (DCP) layer scans, respectively, from human and porcine retinas. For both models, SCP layer was anatomically defined the retinal tissue structure from inner limiting membrane to inner plexiform layer; where as DCP layer was defined as the retinal tissue structure from outer boundary of inner plexiform layer to the outer plexiform layer [52, 53].

**Figure 1.**
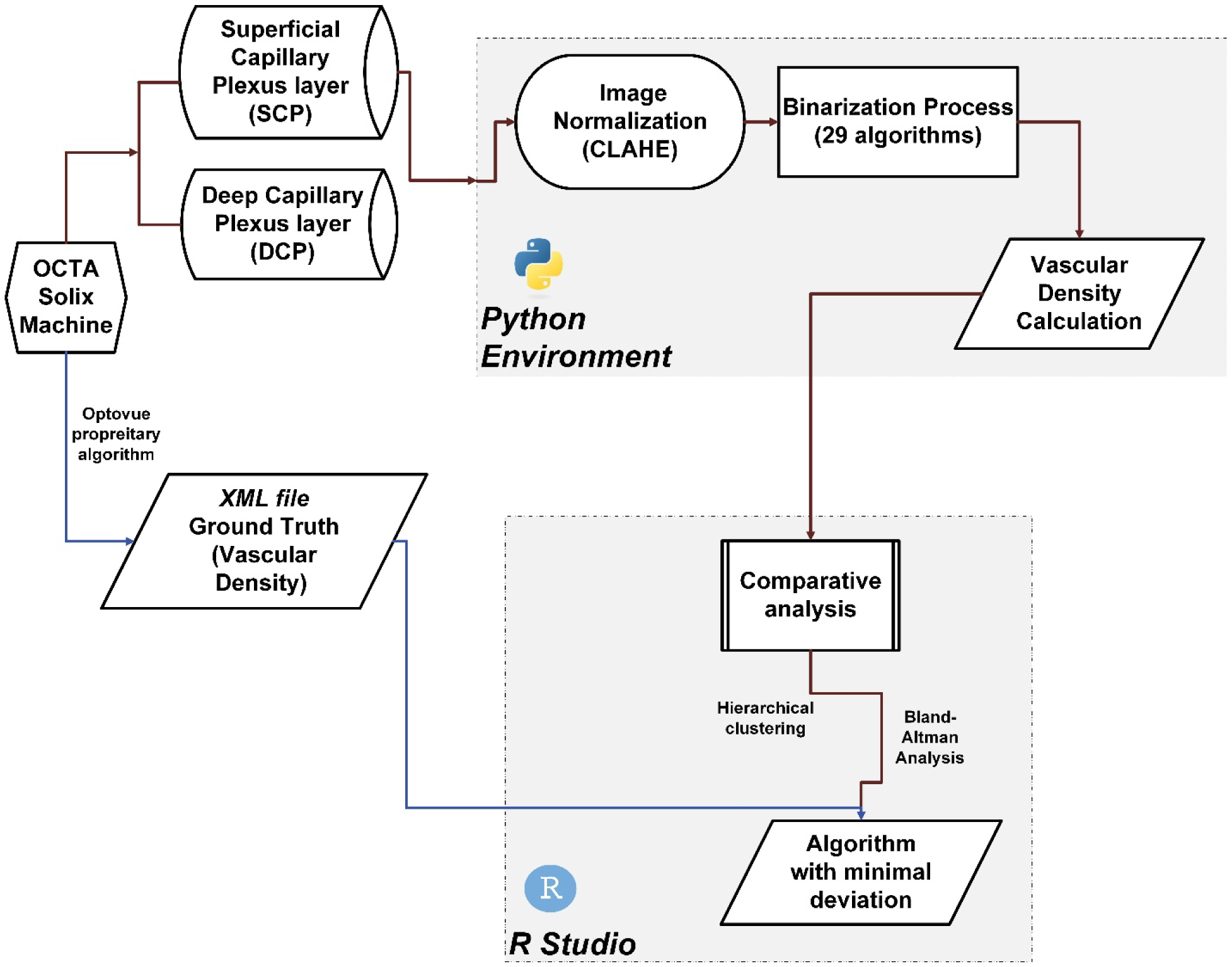
Detailed steps involved in this study are outlined in this flowchart.

Optovue software automatically segments the retinal layers, produces *en face* SCP and DCP scans, and calculates VD for the mentioned *en face* OCTA images (SCP and DCP). OCTA metrics (i.e., VD) for both layers, quantified from the Optovue software, were exported in XML file format. The obtain VD values from Optovue for both models (and respective anatomical layers) served as the “Ground Truth” for analytical purposes.

### Clinical study population

All patient scans were conducted in accordance with University of Texas Southwestern Medical Center Institutional Review Board (IRB) approval. We enrolled patients for OCTA scanning study prior to commencement of surgical procedure in the surgery unit of University of Texas Southwestern Medical Center. The enrolled patients (December 2023 – July 2025) filled out informed consent forms and were not under the influence of anesthesia or any other drugs during the scanning procedure. Inclusion criteria was ≥ 18 years of age and undergoing endoscopic procedures. Patients were excluded if they had any prior surgical procedures in the eye, glaucoma, diabetic retinopathy, retinal detachments, known ocular trauma/injury, ocular tumors, history of contact lenses, pregnancy, etc.

### Clinical Image Acquisition

Clinical OCTA scanning was performed using the FDA approved, Optovue ® Solix acquisition system (Visionix International SAS (formerly Optovue), Pont-de-l’Arche –France) (15 μm/pixel sampling density [54], light source of 840nm; scan speed 120kHz; axial resolution 5μm; lateral resolution 15μm; scan depth up to 3mm; scan width 3-16mm) (JSR, KA, SSP, or UH). Patients were seated in front of the image capture unit and routine OCTA scanning of a single eye was performed (AngioVue Retina mode; 6.4mmx6.4mm scans, 512x512 pixel resolution) to capture the microvascular flow pattern. Ultimately, 17 patients (mean age 63.9 ± 14.4 years, 66.67 % female) were included in the analysis. 22 *en face* OCTA (12 OD scans; 9 OS scans) scans (SCP and DCP layers) were utilized and exported in PNG format for further processing.

### Animal Preparation

Porcine animal scans for this study were obtained as part of our larger ongoing preclinical investigations [6, 50]. Four porcine animals (*Sus scrofa domesticus*, Yorkshire cross; 3 male and 1 female; average weight 31.9 ± 9.8 kg; 2-4 months old) were procured from a commercial vendor (Oak Hill Genetics, Ewing, IL, USA) about 1-2 weeks prior to the experiment. Animals were acclimatized for at least one week in an AAALAC International (Association for Assessment and Accreditation of Laboratory Animal Care) accredited facility at the University of Texas Southwestern Medical Center. Routine diet (Teklad Mini swine Diet 7037, Envigo, Madison, WI) and ad-libitum water were provided to these animals. Room conditions were set to a 12-hour light/dark cycle and standard housing temperature (18-22°C)/humidity (30-70%) settings were maintained. Animals were group or pair housed on slatted or tenderfoot flooring in elevated pens with floor space exceeding the minimum space recommended by the Guide for the Care and Use of Laboratory Animals [55] throughout the study.

Experimental ocular OCTA scans were conducted in accordance protocols approved by the University of Texas Southwestern Institutional Animal Care and Use Committee (IACUC), and we adhered to ARRIVE guidelines (Animal Research: Reporting of In Vivo Experiments) for this investigation. Per previously established protocols [6, 56, 57], animals were sedated by intramuscular injections of telazol 4-6mg/kg followed by glycopyrrolate 0.005 mg/kg as a premedication to reduce bradycardia and salivation. Animals were then placed on a face mask with isoflurane 3-5% to facilitate endotracheal intubation and intravenous and arterial catheter placement. Animals were maintained on 1.5-2% isoflurane and oxygen (2L/min) throughout the experiment to keep the eye in optimal positioning. Mechanical ventilation at 15-20 breath per minute and normal saline (Normasol-R, ICU Medical, Inc., San Clemente, CA, USA) was administered throughout the experiment at a rate of 5-10ml/kg/hr. A heated air blanket (Bair hugger, 3M, St. Paul, MN, USA) was utilized to maintain body temperature and straps were utilized to secure the animal in place for the experiment. Vital signs including arterial blood pressure, end-tidal CO_2_ , heart rate, and pulse were monitored throughout the experiment (SurgiVet Advisor® tech Vital Signs Monitor, Smiths Medical, Inc., Minneapolis, MN, USA). All animals utilized in the study were part of multiple ongoing experiments and not solely for OCTA scanning purposes. Animals from respective studies were euthanized at the end of the experiment with an overdose of pentobarbital sodium (Euthasol®, Virbac, Fort Worth, TX, USA) intravenously.

### Animal Image Acquisition

To facilitate scanning procedure, the animals were placed in prone position on the operating tables and the left eye of the animal was utilized for capturing OCTA images. A speculum was used to keep the eye assessable. Frequent saline flushes were performed to keep the eyes moisturized. OCTA imaging of the porcine eyes was performed using Optovue ® Solix acquisition system (Visionix International SAS (formerly Optovue), Pont-de-l’Arche –France) (15 μm/pixel sampling density [54], light source of 840nm; scan speed 120kHz; axial resolution 5μm; lateral resolution 15μm; scan depth up to 3mm; scan width 3-16mm) (JSR, KA, SSP, or UH). The head of the animal was manually positioned close to image capture unit, and for each acquisition the optimal focal length (prompted by acquisition software) had to be achieved in order to capture the OCTA scan. The animals were draped with black cloth to facilitate hassle-free OCTA acquisitions. OCTA scans of the porcine macular region were performed using the AngioVue Retina mode (6.4mmx6.4mm scans, 512x512 pixel resolution). A total of 29 *en face* OCTA scans were acquired from the four porcine animals. All scans were exported in PNG format for further analysis.

### Image Preprocessing

Automatic, layer-specific segmentation of retinal microvasculature was performed in the AngioVue software. OCTA scans (512x512 pixels; retinal superficial capillary plexus (SCP) and deep capillary plexus (DCP) layers) were excluded if quality was less than 6 on a scale of 10 [58], motion artifacts were present or abnormal vessel segmentation were detected in either retinal layer. Prior to binarization, all images were subjected to contrast-limited adaptive histogram equalization (CLAHE) using FIJI v2.15.1 (National Institutes of Health, available at https://imagej.net/Fiji/Downloads) [59] and a custom Python script to normalize the image intensity. CLAHE was applied with the following parameters: block size of 110 pixels, histogram bin size of 100, and clip limit of 0.01 (adapted from [11, 60]; Supplemental Section S1).

Brightness and contrast were not adjusted to prevent manual bias. Custom Python scripts were utilized in the JupyterLab environment [61] for image analysis process and details are provided in the following sections (GitHub: https://github.com/isi-11/OCTA-Binarization).

### Binarization Techniques

Normalized images were processed using three main approaches: (1) seventeen binarization algorithms in ImageJ (NIH); (2) Random Walker (RW) segmentation; and (3) eleven binarization algorithms from the DoxaPy Python library. The seventeen ImageJ algorithms included: Default (a variation of the IsoData algorithm) [27], Huang [36], Intermodes [24], IsoData [28], Li [38], MaxEntropy [29], Mean [34], MinError [33], Minimum [24], Moments [31], Otsu [25], Percentile [23], Phansalkar [46], RenyiEntropy [29], Shanbhag [35], Triangle [26], and Yen [37]. RW segmentation [41] was implemented in a custom Python script using the random walker function within the scikit-image library [62]. The eleven DoxaPy algorithms included: Bataineh [45], Bernsen [32], Gatos [42], ISauvola [48], Niblack [30], Nick [43], Sauvola [39], Su [44], TRSingh [47], Wan [49], and Wolf [40].

These binarization techniques can be further categorized by the type of thresholding: global or local (adaptive). Global thresholding applies a single threshold value across the entire image; thus, it works best when the image has consistent lighting and contrast. These methods are computationally efficient, but they have been shown to perform poorly in OCTA images with regional intensity variations, often resulting in lower repeatability and less accurate vascular segmentation compared to local thresholding approaches [63]. On the other hand, local thresholding calculates a threshold for each pixel based on its local neighborhood (is specific region or window). This approach is more robust in images with uneven illumination or contrast which is common in OCTA scans. The global thresholding methods evaluated in this study included Default, Huang, Intermodes, Li, MaxEntropy, Mean, Moments, Otsu, Percentile, RenyiEntropy, Shanbhag, Yen, IsoData, MinError, Minimum, and Triangle. The local thresholding included Phansalkar, Niblack, Sauvola, Wolf, ISauvola, Wan, Bernsen, TRSingh, Nick, Su, Gatos, Bataineh, and the Random Walker (RW) segmentation algorithm. Peak Signal-to-Noise Ratio (PSNR) (decibels(dB)) was used to quantify image quality relative to the original grayscale image. Higher PSNR values indicate better preservation of structural details.

### Quantitative Analysis

A total of 22 human and 29 porcine *en face* OCTA scans, respectively, were selected for quantitative analysis. Vascular density (VD) measurements for SCP and DCP layers for each scan were obtained from the exported XML file (Optovue) and parsed through our custom Python script. These values were considered “ground truth” for comparison against the VD from the binarized images. Additionally, unlike humans, pigs do not have a foveal avascular zone (FAZ) [6], thus only VD quantification metric from the Optovue software were utilized for comparative purposes. Next, for both the SCP and DCP binarized images, vascular density (VD) was computed as the percentage of the image area occupied by binarized vessel pixels (total number of white pixels over total pixel in the entire image). These calculated VD values for each algorithm were then compared against Optovue-reported VD values (“ground truth”).

### Statistical Analysis

All statistical analyses were performed using GraphPad® Prism 10 and R Studio [64] - *ggplot2 [65], dplyr [66], gridExtra [67], factoextra [68], cluster [69],* and *tidyr [70]* packages. The continuous variable, VD, was reported as mean ± standard deviation (%). Pairwise comparison between Optovue and each of the 29 algorithm laden VD values were compared using paired t test and Benjamini-Hochberg correction was applied (*q*-value) to avoid false discovery rate (α = 0.05). We analyzed whether global and local algorithms tend to form distinct functional clusters via hierarchical clustering methodology. Quantified VD data was scaled and the optimal number of clusters were determined using the Silhouette method [71]. After the optimal number of clusters were obtained, we performed hierarchical clustering with Euclidean distance metrics and complete linkage was then used to generate dendrograms separately for each retinal layer of human and porcine cohorts, visualizing the grouping of algorithms. Further, to evaluate systemic bias and agreement between each binarization algorithm and Optovue reference values (ground truth), Bland-Altman analyses were conducted for each layer for both models.

The outcome of the Bland-Altman analyses was reported as the mean difference and 95% limits of agreement for each method (plots generated using BA-plotteR) [72].

## Results

### Method-Dependent Variability in OCTA Image Binarization

OCTA images were subjected to adaptive, localized CLAHE equalization, followed by the 29 binarization algorithms, and then then VD and PSNR metrics were quantified (details of PSNR quantification can be found in Supplementary Table S2). Exemplary steps in binarization of OCTA scan from human and porcine models, respectively, for the SCP layer is shown here in Fig. 2 (A-G).

**Figure 2.**
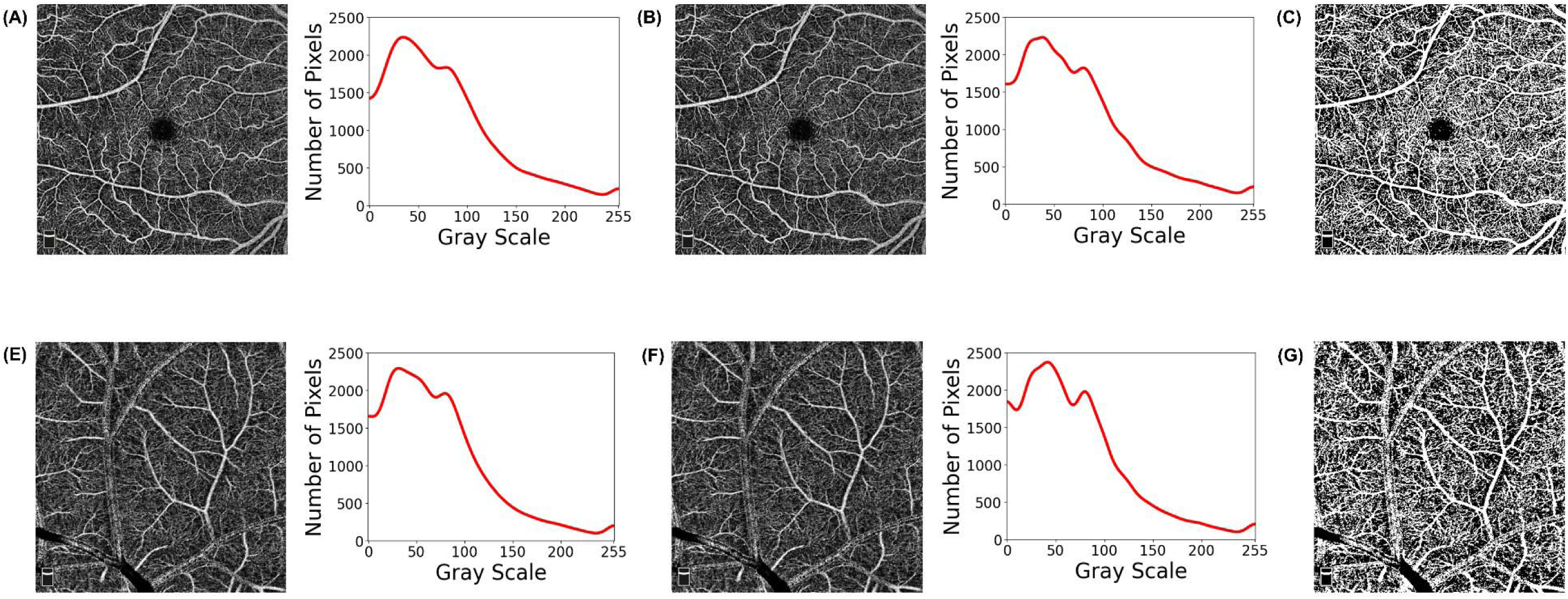
OCTA image processing steps involved in this study are shown here. Exemplary OCTA scans of human (A) and porcine (E) subjects, respectively, are shown here. Subsequent image normalization using Contrast-Limited Adaptive Histogram Equalization (CLAHE) ((B) & (F)), and subsequent binarized output ((C) Sauvola & (G) Wolf) is provided here as well.

Qualitative comparison of the original and algorithm-laden binarized versions for human SCP, human DCP, porcine SCP, and porcine DCP layers are shown in Figs. 3(a-d), respectively. For the human cohort, the PSNR for both SCP and DCP layers was found to be highest for Bernsen (11.1 ± 0.3 dB for SCP; 10.3 ± 0.3 dB for DCP) and lowest for Su (2.3 ± 0.1 dB for SCP; 2.7 ± 0.2 dB), respectively (Fig.3(a)). Magnitude of PSNR for human DCP scans when Moments algorithm was applied, were similar to Su algorithm; however, vascular structures were better preserved in Moments algorithm (Fig. 3(b)). Similar pattern was observed in the porcine cohort as well, and the Bernsen algorithm produced the highest PSNR for the SCP and DCP layers, respectively (11.4 ± 0.4 dB for SCP; 10.8 ± 0.4 dB for DCP). The lowest PNSR magnitude for porcine SCP layer was Su algorithm (2.2 ± 0.1 dB) like the human cohort (Fig. 3(c)). However, the PNSR for porcine DCP layer was similar in magnitude across MaxEntropy, Renyi Entropy, Su, and Yen algorithms (2.4 ± 0.2 dB) and Su algorithm exhibited better visual quality as compared the others (Fig. 3(d)). Highest PSNR magnitudes belonged to the localized thresholding type (Bernsen), whereas the lower PSNR ones were primarily global thresholding type (Moments, RenyiEntropy, and Yen algorithms) with the exception of Su (localized thresholding).

**Figure 3(a).**
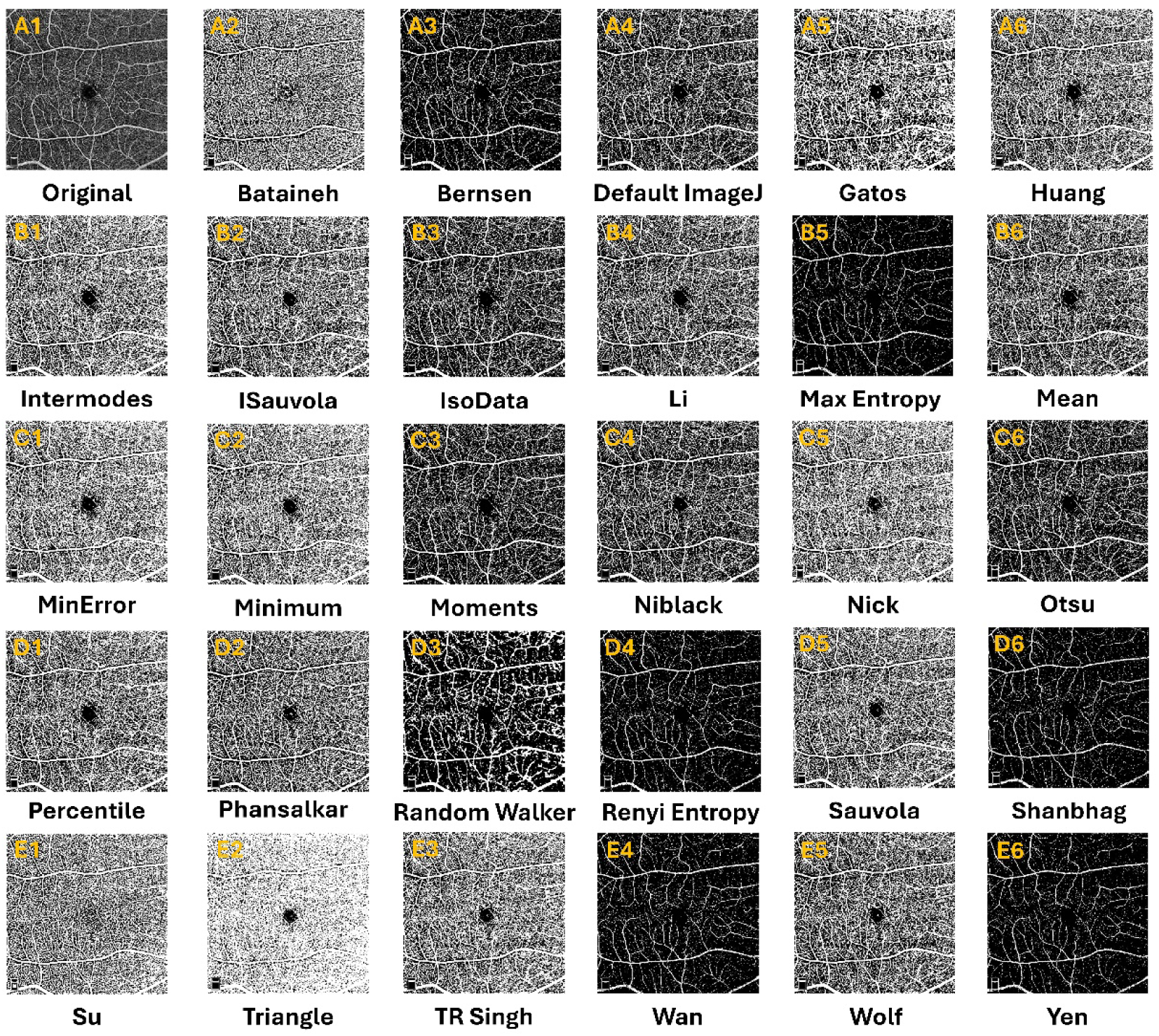
The impact of 29 unique binarization algorithms on human OCTA scans – superficial capillary plexus (SCP) - is shown here. *First row*: (A1) Original OCTA scan; (A2-A6) Bataineh, Bernsen, ImageJ Default, Gatos, and Huang. *Second Row*: (B1-B6) Intermodes, ISauvola, IsoData, Li, MaxEntropy, and Mean. *Third Row* (C1-C6): MinError, Minimum, Moments, Niblack, Nick, and Otsu. *Fourth Row* (D1-D6): Percentile, Phansalkar, RandomWalker, RenyiEntropy, Sauvola, and Shanbhag. *Fifth Row* (E1-E6): Su, Triangle, TRSingh, Wan, Wolf and Yen.

**Figure 3(b).**
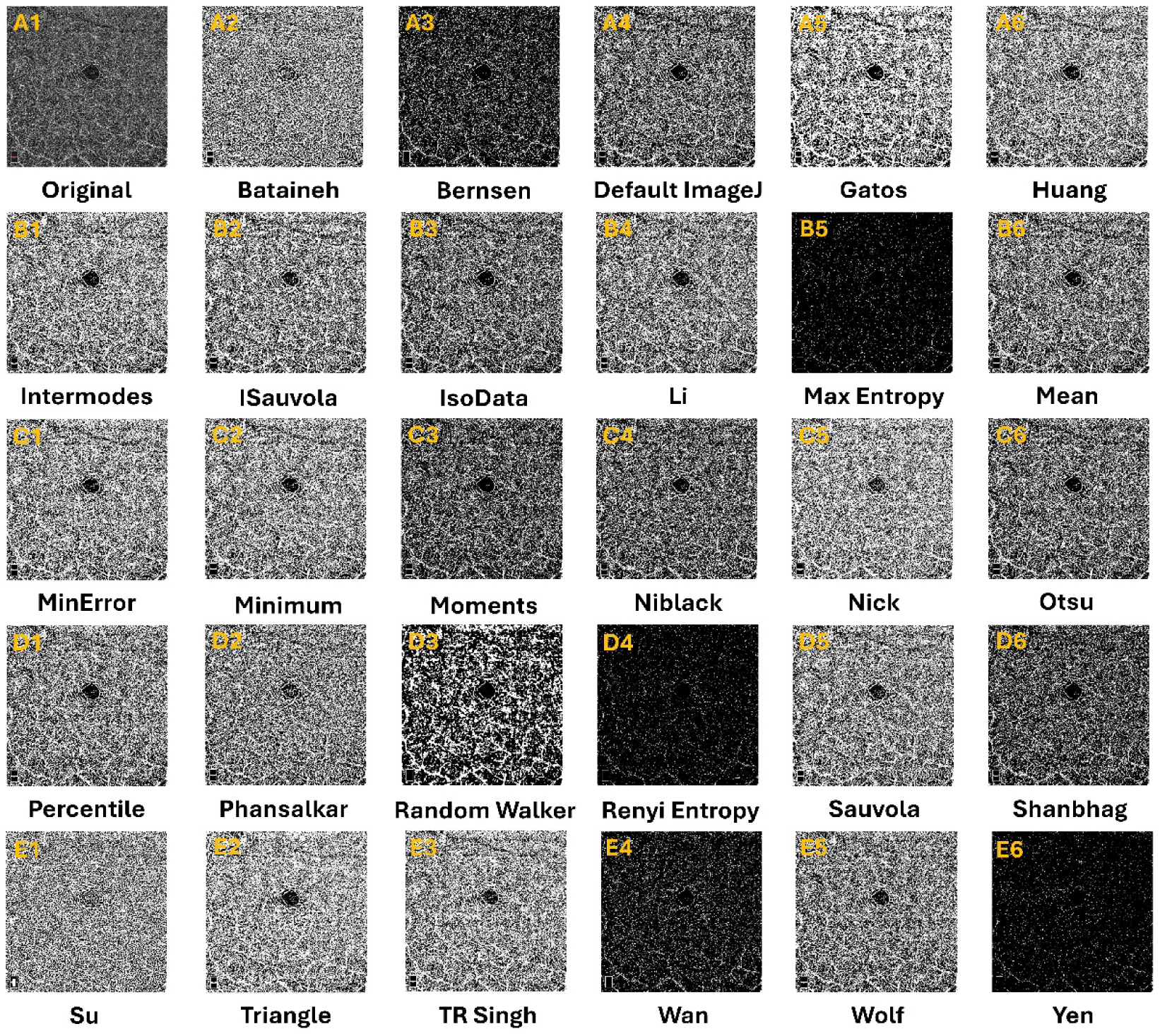
The impact of 29 unique binarization algorithms on human OCTA scans – deep capillary plexus (DCP) - is shown here. *First row*: (A1) Original OCTA scan; (A2-A6) Bataineh, Bernsen, ImageJ Default, Gatos, and Huang. *Second Row*: (B1-B6) Intermodes, ISauvola, IsoData, Li, MaxEntropy, and Mean. *Third Row* (C1-C6): MinError, Minimum, Moments, Niblack, Nick, and Otsu. *Fourth Row* (D1-D6): Percentile, Phansalkar, RandomWalker, RenyiEntropy, Sauvola, and Shanbhag. *Fifth Row* (E1-E6): Su, Triangle, TRSingh, Wan, Wolf and Yen.

**Figure 3(c).**
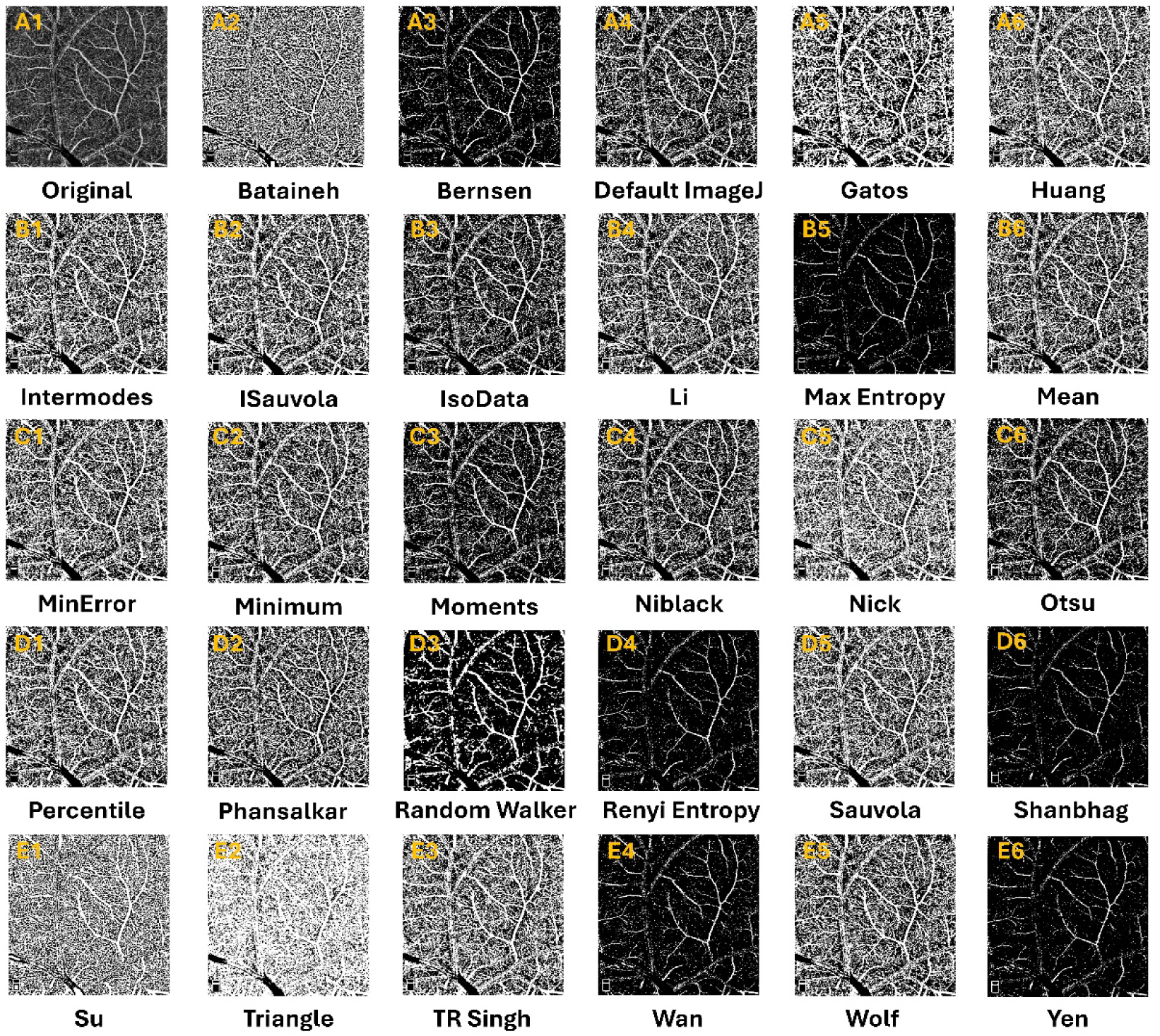
The impact of 29 unique binarization algorithms on porcine OCTA scans – superficial capillary plexus (SCP) - is shown here. *First row*: (A1) Original OCTA scan; (A2-A6) Bataineh, Bernsen, ImageJ Default, Gatos, and Huang. *Second Row*: (B1-B6) Intermodes, ISauvola, IsoData, Li, MaxEntropy, and Mean. *Third Row* (C1-C6): MinError, Minimum, Moments, Niblack, Nick, and Otsu. *Fourth Row* (D1-D6): Percentile, Phansalkar, RandomWalker, RenyiEntropy, Sauvola, and Shanbhag. *Fifth Row* (E1-E6): Su, Triangle, TRSingh, Wan, Wolf and Yen.

**Figure 3(d).**
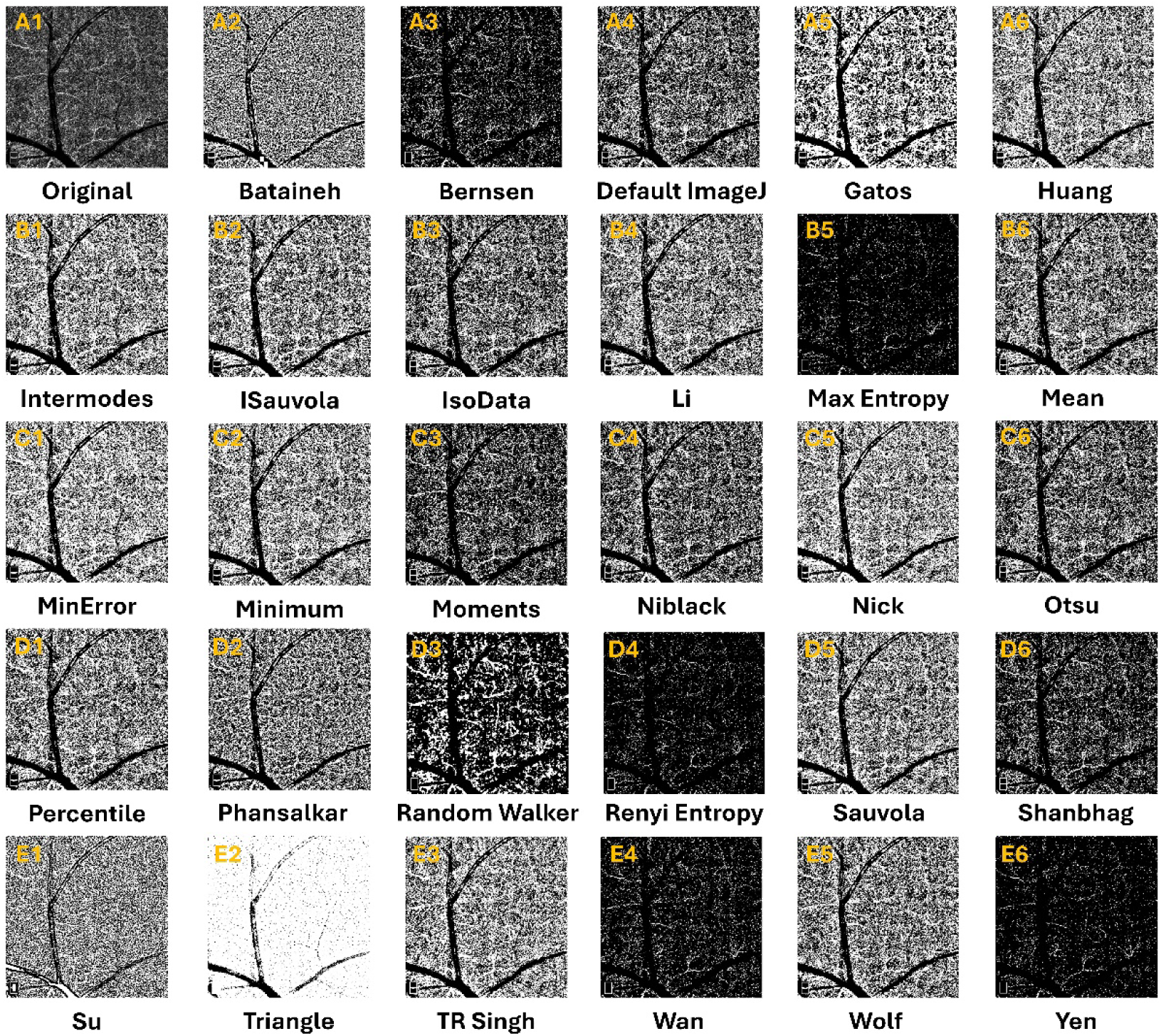
The impact of 29 unique binarization algorithms on porcine OCTA scans – deep capillary plexus (DCP) - is shown here. *First row*: (A1) Original OCTA scan; (A2-A6) Bataineh, Bernsen, ImageJ Default, Gatos, and Huang. *Second Row*: (B1-B6) Intermodes, ISauvola, IsoData, Li, MaxEntropy, and Mean. *Third Row* (C1-C6): MinError, Minimum, Moments, Niblack, Nick, and Otsu. *Fourth Row* (D1-D6): Percentile, Phansalkar, RandomWalker, RenyiEntropy, Sauvola, and Shanbhag. *Fifth Row* (E1-E6): Su, Triangle, TRSingh, Wan, Wolf and Yen.

### Inter-Method Comparison of Vascular Densities

Detailed vascular densities of original and their algorithm-laden binarized versions of OCTA scans for human SCP and DCP, and porcine SCP and DCP layers, respectively, are shown in Table 1 (a) (Supplemental section S3). VD values varied substantially across binarization algorithms for both human and porcine OCTA datasets. In human scans, SCP VD ranged from 10.7±1.0% (Wan) to 91.4±1.1% (Yen), while DCP VD ranged from 11.3±1.3% (Wan) to 94.3±1.2% (Yen). Similar trends were observed in porcine scans, with SCP VD spanning 9.8±0.9% (Wan) to 91.8±1.3 (Yen)% and DCP VD from 10.0±1.0% (Wan) to 94.3±0.9%(Yen).

Algorithms utilizing entropy-based global thresholding (MaxEntropy, RenyiEntropy, Yen) consistently produced the highest VD values (>90%), whereas local thresholding methods such as Bernsen and Wan yielded the lowest (<20%). Optovue’s native algorithm reported intermediate VD values (SCP: 48.3±1.4% and DCP: 51.9±1.7%, respectively in humans; SCP: 46.2±1.8% and DCP: 50.0±1.4%, respectively, in porcine).

For the SCP layer in human scans (Table 1 (b)), most algorithms demonstrated statistically significant differences compared to Optovue (46.2±1.8%), except Huang (48.5 ± 5.2%; *p*=0.83) and Minimum (47.2 ± 8.1%; *p*=0.55). The largest deviations were observed for Wan (10.7 ± 1.0%; t=140.95, *p*=9.64 x 10^-33^) and MaxEntropy (91.1 ± 1.0%; t=-109.99, *p*=1.75x 10^-30^), indicating extreme underestimation and overestimation, respectively, relative to Optovue. Similar patterns were noted in the DCP layer (Table 1 (c)), where all algorithms except Random Walker (54.2 ± 8.5%; *p*=0.14) did not differ significantly. Entropy-based global thresholding methods again showed the greatest positive divergence (e.g., Yen: t=-79.36, *p*=1.63E-27) while Bernsen and Wan showed the greatest negative divergence (e.g., Wan: t=213.52, *p*=1.58E-36).

For the SCP layer in porcine scans (Tables 1 (d)), most binarization algorithms produced VD values significantly different from Optovue (46.2 ± 1.8%). Exceptions included Huang (46.9 ± 4.8%; *p*=0.47), Intermodes (50.3 ± 1.5%; *p*=0.09), Minimum (48.4 ± 5.9%; *p*=0.07), and Wolf (46.3 ± 1.4%; *p*=0.74), which were not statistically different from Optovue. All other algorithms showed significant differences, with entropy-based global thresholding methods (e.g., MaxEntropy, Renyi Entropy, Yen) yielding the largest overestimations (e.g., Yen: 91.8 ± 1.3%; t=-143.29, *p*=1.13x 10^-41^) and Bernsen and Wan producing the largest underestimations (e.g., Wan: 9.8 ± 0.9%; t=141.24, *p*=1.69 x 10^-41^). Similar trends were observed in the DCP layer (Table 1 (e)), where nearly all algorithms differed significantly from Optovue (50.0 ± 1.4%), except Li (50.5 ± 2.4%; *p*=0.23) and Percentile (50.1 ± 1.5%; *p*=0.75). Again, global entropy-based methods produced extreme VD overestimations compared to Optovue (e.g., Yen: 94.3 ± 0.8%; t=-166.54, *p*=1.69x 10^-43^) while Bernsen and Wan produced extreme underestimations (e.g., Wan: 10.0 ± 1.0%; t=146.87, *p*=5.66x 10^-42^).

### Hierarchical Clustering of Estimated Vascular Densities from 29 algorithms

Unsupervised clustering was performed to evaluate similarity among binarization algorithms based on VD outputs for SCP and DCP layers in both human and porcine OCTA scans, respectively. For the SCP layer, the Silhouette method indicated that three clusters were optimal for the human cohort (Fig.4A), whereas four clusters were optimal for the porcine cohort (Fig.4B). Dendrograms revealed distinct grouping patterns, with entropy-based global thresholding algorithms (e.g., MaxEntropy, Renyi, Yen) clustering together due to their consistently high VD values, while local adaptive methods (e.g., Bernsen, Wan) formed separate clusters characterized by low VD outputs (Fig.4C-D). The largest cluster for human SCP layer was formed by Minimum, Intermodes, Random Walker, MinError, Wolf, Phansalkar, Triangle, Niblack, Sauvola, iSauvola, Gatos, Percentile, Huang, Mean, Li, Su, Nick, TR Singh, and Bataineh algorithms (almost 50% between global and local thresholding categories), and the smallest cluster was formed by Wan and Bernsen (localized thresholding). On the other hand, the largest cluster in the porcine SCP cohort constituted of Sauvola, iSauvola, Gatos, Percentile, TR Singh, Bataineh, Su, Nick, Mean, Li, Wolf, Phansalkar, Huang, Minimum, Intermodes, and MinError – very similar to their human counterpart. Further, two smallest clusters were formed for the porcine SCP – (1) Otsu, Moments, IsoData, and Default ImageJ, and (2) Renyi Entropy, Max Entropy, Yen, and Shanbhag – both smaller clusters belong to global thresholding type.

**Figure 4.**
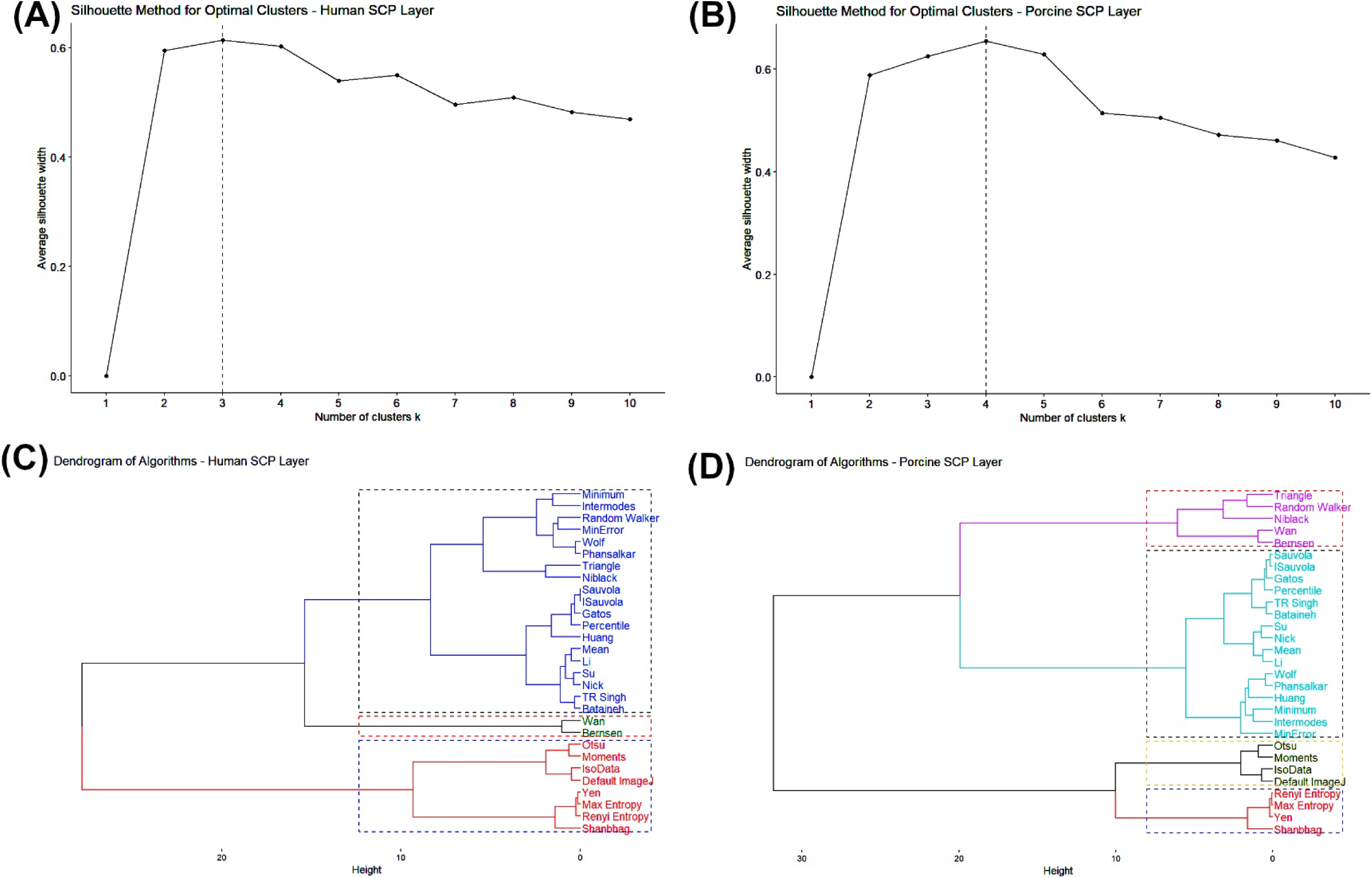
Clustering analysis of vascular density metrics from the investigated algorithms for the human and porcine SCP layers, respectively, are reported here. Silhouette method showed that three clusters are optimal for human cohort and four clusters for porcine cohorts, respectively (A-B). Dendrograms of both cohorts show the similarity between algorithms in human (C) and porcine (D) clusters, respectively, for SCP vascular density metrics.

For the DCP layer, the Silhouette method identified three optimal clusters for the human cohort (Fig.5A), similar to the SCP, and three clusters for the porcine cohort (Fig.5B). The DCP showed similar groupings in regard to the entropy-based global thresholding algorithms and Bernsen and Wan as the SCP (Fig.5C, Fig.5D). Dendrograms for both cohorts showed heterogeneity in algorithm grouping for DCP, with distinct clusters reflecting variability in VD estimation. For Human DCP cohort, the largest cluster contained Sauvola, iSauvola, Bataineh, Mean, Wolf, Percentile, Phansalkar, Li, Random Walker, Minimum, Intermodes, MinError, Huang, Triangle, Niblack, TR Singh, Su, IsoData, Default ImageJ, Otsu, Nick, Gatos, Shanbhag, and Moments (11 localized type out of 24 total). Wan, Bernsen, and Triangle algorithms formed the smallest cluster in the porcine DCP cohort (Triangle belongs to the global thresholding type).

**Figure 5.**
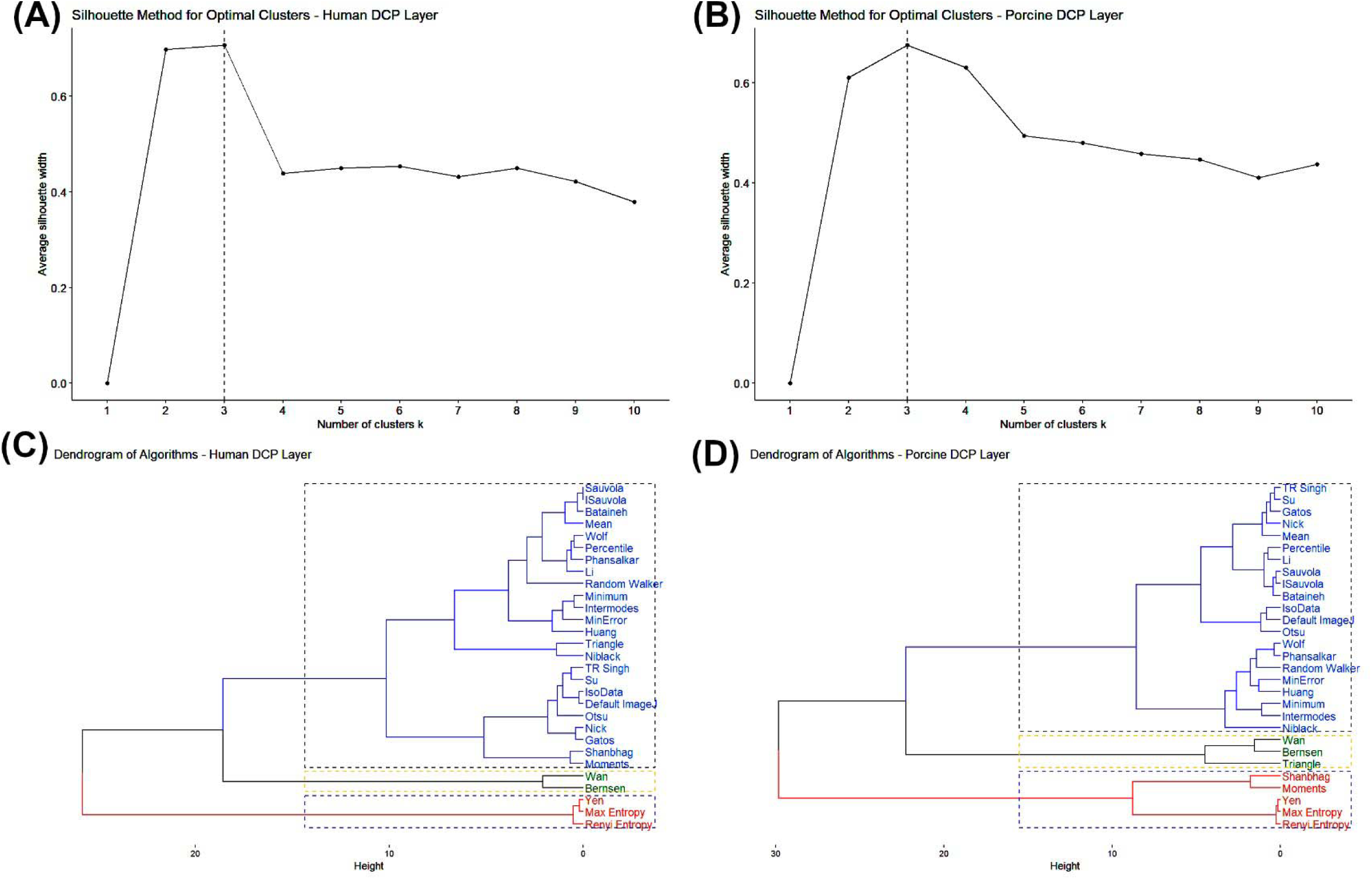
Clustering analysis of vascular density metrics from the investigated algorithms for the human and porcine DCP layers, respectively, are reported here. Like SCP layer, the Silhouette method showed that three clusters are optimal for DCP layer from the human cohort (A). However, only three clusters were found to be optimal for DCP layer from porcine cohort (B). Dendrograms of both cohorts show the heterogeneity between algorithms in human (C) and porcine (D) clusters, respectively, for DCP vascular density metrics.

### Agreement between the two methods – Ground Truth vs. Binarized algorithms

Bland-Altman analysis for the human SCP layer revealed that that Yen algorithm exhibited a large, estimated bias (mean difference) 43.05%(95% CI : 42.21% to 43.9%) (CI= confidence interval) as compared to Optovue, and the least deviation estimated bias from ground truth (Optovue) was for Wolf algorithm – difference of -1.78% (95% CI: -2.5% to -1.06%) (Fig.6 (A1-A2)). For porcine SCP, the algorithm with maximum bias was Shanbhag algorithm – mean difference 44.26% (95% CI: 40.66% to 47.86%) (Fig.6 (A3)). Similar to human SCP, Wolf algorithm also exhibited least bias in comparison to porcine SCP Optovue values – mean difference of 0.09% (95% CI: -0.54% to 0.72%) (Fig.6 (A4)). Thus, indicating that local thresholding algorithms (Wolf) have superior performance over global thresholding algorithms (Yen and Shanbhag) for SCP layer from both models.

**Figure 6.**
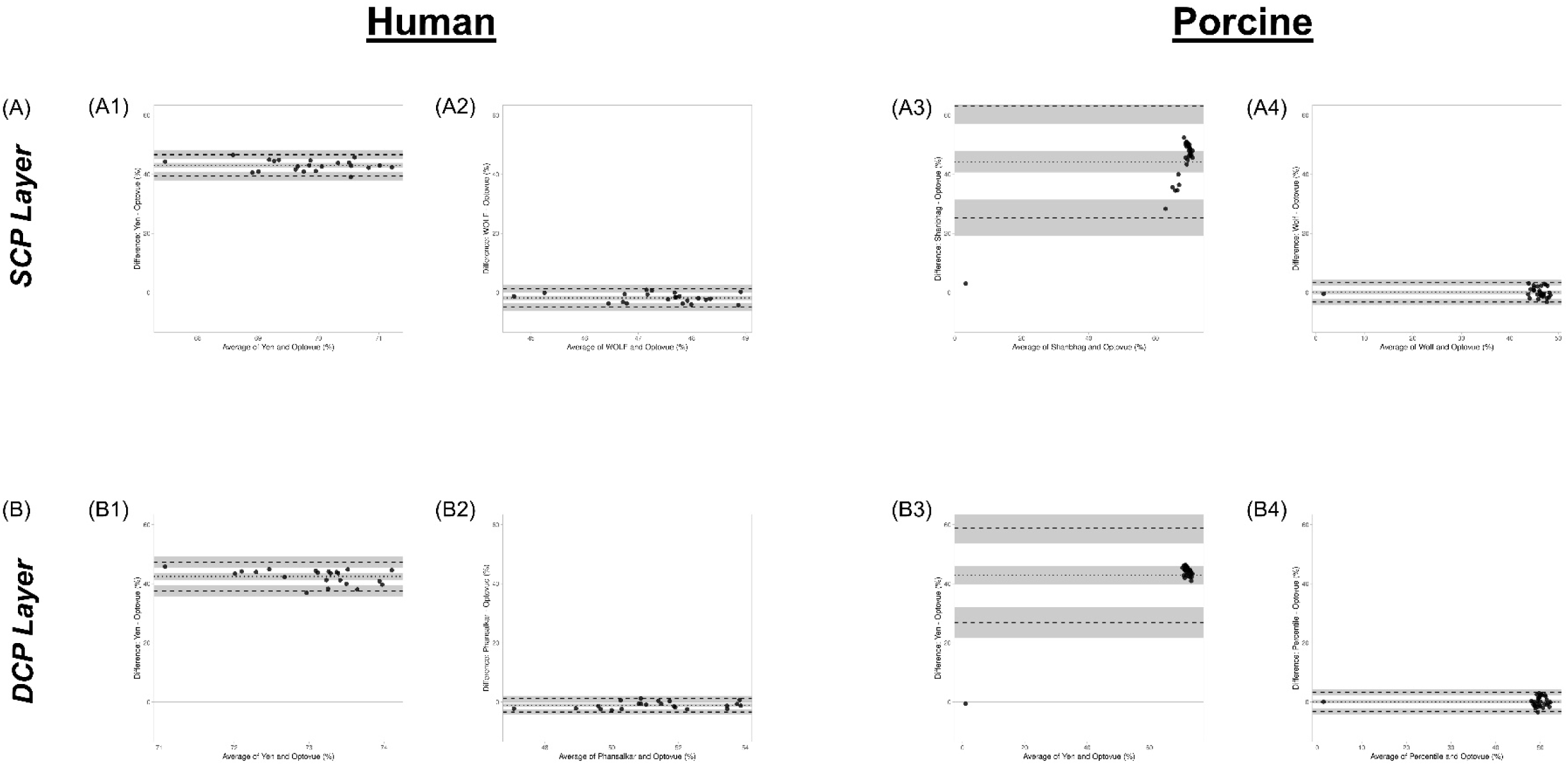
Bland-Altman analysis of vascular density (VD) obtained from Optovue (ground truth) compared to 29 binarization algorithms (expressed in %). For the human SCP layer, Yen and Wolf exhibited the worst and best comparison to Optovue, respectively (A1-A2). Wolf was the best for porcine SCP as well, and on the other hand, Shanbhag was the worst (A3-A4). Yen did not perform well for both human and porcine DCP layers, respectively (B1 & B3). Phansalkar and Percentile algorithms exhibited minimal, estimated bias from Optovue values, respectively, for human and porcine DCP layers (B2 & B4). Each plot includes mean difference (bias) (dotted line) and 95% limits of agreement (dashed lines and grey boundaries).

Similar to human SCP layer, Yen algorithm exhibited highest estimated bias (mean difference) for human DCP layer from Optovue DCP values as well – mean difference of 42.45% (95%CI: 41.31% to 43.59%) (Fig. 6(B1)). Phansalkar algorithm on the other hand exhibited minimal, estimated bias from ground truth VD values – mean difference of -1.11% (95%CI: -1.64% to -0.58%) (Fig.6(B2)). Again, for the porcine DCP layer, the Yen algorithm exhibited a substantial, mean difference of 42.84% (95%CI: 39.8% to 45.89%) (Fig.6(B3)). The least bias for porcine DCP layer was produced by Percentile algorithm (mean difference of 0.1 (95%CI: -0.52% to 0.72%) (Fig. 6(B4)). Phansalkar algorithm (localized type) produced the least deviation from ground truth for human DCP cohort. Interestingly, Percentile (global type) exhibited the least deviation from ground truth for porcine DCP cohort.

## Discussion

An OCTA scan indicates the presence or absence of retinal blood flow (not direct quantitative metric), and the vascular density metric utilized for clinical diagnostic purposes is a derived, semi-quantitative biomarker. Perfusion status of the retinal vasculature can function as a surrogate for cerebral microcirculation and this biomarker is particularly useful for severely ill patients [3]. Now, the challenge is the interpretation of the OCTA scans. OCTA scans’ perfusion status is provided by a multitude of biomarkers [3, 7, 8, 22] and most common being the vascular density metric. Currently, every OCTA manufacturer has its own proprietary quantification/binarization/segmentation algorithm for calculating OCTA biomarkers (including vascular densities) and hence, the VD metric varies across acquisition units. Further, quantitative OCTA metrics, especially VD, obtained from clinical patients have shown to vary between hardware system [15, 18] or software versions [73], and have demonstrated reproducibility problem [21, 74, 75]. Although several binarization techniques have been explored in digital image processing, relatively few have been systematically evaluated in the context of OCTA [11, 12]. If we perform offline analysis, like our study, the blending in image processing steps for VD quantification are endless. To make the OCTA biomarkers robust and repeatable, there is a dire need for standardization of scanning and quantification procedures.

Our goal from this study was to unravel the influence of the critical step of binarization in the quantification of vascular density metric from OCTA scans. We evaluated the binarization techniques (after normalization with CLAHE) using PSNR, which provided pixel-wise quantitative changes in the image after binarization algorithm was applied. Next, we compared the average quantitative values from the Optovue software with the calculated VD from the binarized images, and this analysis provided an insight to algorithms that were really close or far to VD values produced from the source. After that we evaluated which algorithms produced similar VD values when they were clustered together. Lastly, we complemented the aforementioned analysis by finding which techniques produced minimal changes from the mean values obtained from Optovue software. Our findings suggest that (i) binarization algorithms are varied and not interchangeable, (ii) no single algorithm is best fit for either retinal tissue layer vascular density quantification (collectively, local threshold methods are superior to global ones), (iii) SCP layer is more vulnerable to binarization algorithms, and (iv) DCP layer is less susceptible to binarization algorithms.

### No Universal Binarization Algorithm for Diverse Retinal Tissues

There is no direct consensus for the image processing steps utilized for analysis of OCTA scans. Prior studies have shown that use of CLAHE or background subtraction pre-processing steps may offer a potential standardization technique and it is helpful to reduce the vulnerability associated with the outcomes with the applied binarization technique(s) [76]. As observed in our study (Fig.3(a-d)) and suggested by Tan et al. [77], the appearance of retinal vessels is highly dependent on the type of binarization used and the variability in the processing method across companies make it even harder to reproduce (or difficult to interpret).

There are a only a handful of porcine OCTA studies published in literature to perform a thorough comparative analysis [6, 78–80]; hence, making it difficult to analytically evaluate the influence of binarization on porcine retinal layers (unlike the human ones). Comparison of vascular densities for human cohort is challenging as well, due to the significant variability across devices, retinal layer definitions, angiocube size, signal strength, and other limits [10, 12, 13, 18, 21, 76, 81–83]. Optovue generated vascular density measurements for human subjects in our study were found to be greater in magnitude for DCP layer as compared to the SCP layer (Table 1(a); 51.9 ± 1.7% vs. 48.3 ± 1.4%). This observation is consistent with previously published studies with Optovue devices [10, 81, 84–87] and several other studies [88–90] that report the reverse relationship between SCP and DCP vascular densities.

Our unique contribution from this study was the in-depth analysis of the effect of binarization techniques on quantitative vascular density measurement from human and porcine retinas, and across both SCP & DCP layers. Further, this study is also one of the first to report the analytical effect of global vs. local thresholding on porcine retinal vascular density quantification. Wan, Bernsen, Wolf and Phansalkar algorithms (localized threshold type) dominated the binarization performance across the retinal layers as well as both the models; Percentile algorithm performance was an exception for porcine DCP cohort (Figs. 3-6).

The superior performance of local binarization algorithms may be attributed to their adaptive approach compared to global methods. Global techniques apply a single threshold across the entire image, making them highly susceptible to uneven illumination and contrast variations common in OCTA scans [91]. This often leads to vessel dropout, fragmentation, and loss of fine capillaries, particularly in SCP images (due to presence of wide array of vessel diameters as compared to the DCP layer). In contrast, local adaptive algorithms such as Sauvola and Wolf determine thresholds based on neighborhood statistics, enabling them to adjust to regional intensity differences (Fig. 2). This adaptability preserves vessel continuity and microvascular architecture, which is reflected in higher PSNR values and more accurate VD measurements. Prior studies, including those by Arrigo et al.[12] and Laiginhas et al.[63], have emphasized the advantages of local strategies for OCTA quantification, especially in layers with heterogeneous intensity profiles. Our findings build on this evidence by demonstrating these benefits consistently in both human and porcine OCTA images.

Our findings align with prior reports highlighting variability and poor repeatability of OCTA metrics across thresholding strategies [17, 74, 82, 92]. Our interclass correlation (ICC) analysis revealed (data not reported here; supplemental section S4(a-d)) that none of the binarization algorithms exhibited “strong” (ICC of 0.9 or greater) or “excellent” (ICC of 0.75-0.9) associations with the Optovue (ground truth) for either models/layers. Only Phansalkar algorithm (local methods) exhibited “good” and “fair” correlations with Optovue values for human deep capillary plexus (ρ = 0.653) and porcine superficial capillary plexus (ρ = 0.557), respectively. Thereby indicating no fixed pattern in the VD quantification. Arrigo et al. [12] compared 13 thresholding methods and found Huang, Li, Mean, and Percentile to be most reliable, while Intermodes, MaxEntropy, RenyiEntropy, and Yen performed poorly (global methods). Mehta et al. [11] evaluated 11 methods and reported low repeatability (ICC < 0.8) across devices and plexuses, noting that local thresholds sometimes misclassified the FAZ. Corvi et al.[74] demonstrated significant differences in VD across five global thresholds (Default, Huang, IsoData, Mean, Otsu), reinforcing that algorithm choice impacts quantitative metrics. Terheyden et al. [17] on the other hand suggest that automated binarization algorithms in ImageJ (Huang, Li, Otsu, Moments, Mean, and Percentile) exhibited excellent reproducibility of OCTA parameters for their patient cohort, but these algorithms are not interchangeable or readily comparable with one another.

Clearly, the reported data is very diverse (Table 1(a-d)), and no single algorithm is perfect/universal for either retinal layer from humans or porcine sources. Thus, in agreement with published literature [17, 74, 82, 92], we found that the binarization algorithms are not interchangeable and (collectively) that localized, adaptive type thresholding algorithms perform better as compared to a global one. A comparative analysis of the influence of binarization techniques on OCTA metrics has been provided in Table 2.

### SCP is most affected by binarization techniques

Our study revealed that the SCP layer exhibited the greatest variability in VD across binarization algorithms (Table 1(a)). The SCP processed with global thresholds often showed vessel dropout or excessive segmentation, while local adaptive methods preserved microvascular detail and continuity (Fig. 3a & 3c). Human SCP VD differences were most pronounced for global algorithms such as Yen, which overestimated VD by nearly average of 43.05% compared to Optovue’s (Fig.6). The Wolf algorithm on the other hand demonstrated minimal deviation for both human and porcine cohorts, with roughly a 0.09% deviation from mean, respectively. Thus, resulting in the localized thresholding methods the most reliable ones for SCP scans. These findings are consistent with prior literature. Corvi et al. [74] demonstrated significant VD differences across global thresholds in SCP, reinforcing its sensitivity. Laiginhas et al. [63] emphasized the superiority of local strategies for quantification, which parallels our conclusion that local adaptive methods should be prioritized for OCTA image processing. Arrigo et al. [12] reported that SCP metrics were particularly vulnerable to thresholding variability, although they found that global methods primarily underestimated VD. Additionally, Borrelli et al. [91] noted that global thresholds yielded better visual quality, but local thresholds tended to inflate VD values for SCP images. Unlike Borrelli et al. [91], our analysis found that local thresholds generally produced SCP VD values closer to Optovue benchmarks rather than inflating them.

Hence, the effect of binarization on the SCP layer shows conflicting evidence in literature, and our study suggests that SCP scans are more likely to demonstrate variability in vascular density upon binarization (regardless of local or global type).

### DCP is least affected by binarization techniques – our novel contribution

In contrast to the SCP, the DCP layer was relatively robust to binarization variability: a novel finding of this study. VD values for the DCP remained closer to Optovue benchmarks across most algorithms, with narrower variability compared to SCP. For porcine scans, Wolf (local method) produced VD values nearly identical to Optovue, while even less optimal algorithms exhibited smaller deviations than those observed in SCP. As an exception, Percentile algorithm exhibited minimal deviation for the porcine DCP cohort. This observation may be attributed to the difference in anatomical structure between two species (Figs. 2-3). For example, (i) no FAZ is present in porcine retina, and (ii) presence of multiple cilioretinal arteries rather than a single central retinal artery in humans [6, 78]. Our results highlight the importance of layer-specific considerations: while the SCP demands careful algorithm selection, the DCP may tolerate greater flexibility without compromising metric accuracy. Previous studies rarely addressed this layer-specific difference. Mehta et al.[11] noted that deeper layers generally showed higher repeatability than SCP, which aligns with our observation of DCP’s resilience. However, most prior work focused on SCP or full-thickness slabs, making our contribution unique in systematically comparing SCP and DCP sensitivity to binarization.

### Limitations

We acknowledge that this study is not without its limitations. The study’s limitations include modest sample size, single-device acquisition (Optovue), and exclusion of pathological eyes. Signal quality varies between animal and human scans, and this has direct implications on VD calculations (irrespective of binarization technique used) [19, 58, 82, 94]. We utilized all human eyes independently and did not explore the effect of side (OD or OS differences). For animal scans, the right eye was not accessible and hence, we only utilized the left eye for all our experiments. In addition to global and local thresholding algorithms, there is also complex thresholding (combination of global and local binarization methods) techniques [91] that we did not consider for our study. We focused primarily on the VD calculation procedure for this study, whereas other quantitative metrics such as vessel skeletal density, vessel length, vessel area density, etc., could have been explored as well. Although local, adaptive algorithms were superior in their performance as compared to the global ones, it is possible that sometimes the former ones tend to overestimate the vascular density of the image by incorrectly binarizing the foveal avascular zones [11]. Next, the binarization threshold of OCTA scans can also produce artifacts and physiological interpretation of such an anomaly can be complicated [96]. In certain situations, for patients with diabetic retinopathy, it was found that global algorithms perform better than the local ones in terms of qualitative image quality assessment [94]. The same study [94] also found that image quality and presence of macular edema were responsible for overestimation of vascular density metrics (especially, vascular density from the local binarization algorithms). These highlighted limitations underscore the need for harmonization of OCTA image processing techniques and biomarker quantification strategies for vigorous clinical efficiency.

### Recommendation

Clinical utility of OCTA scanning is gaining popularity across disciplines, but a simple question remains unanswered - “What magnitude of change in retinal perfusion status is clinically relevant?”. Before attempting to answer this fundamental query, a simple and harmonized protocol needs to be established for comparing/contrasting OCTA biomarker data across studies or patient populations. To start with a small caveat, we attempted to focus on the widely utilized biomarker – vascular density. For obtaining vascular density from OCTA scans, binarization step is the most fundamental step in OCTA biomarker quantification process. This single critical processing step can either overestimate or underestimate the actual physiological observation. Hence, for better reproducibility and harmonization of OCTA image processing steps, we can provide the following recommendations:

- A research study involving OCTA scanning should report the following

`o` OCTA image acquisition machine details
`o` Software version (if applicable)
`o` Retinal layer to be evaluated
`o` Scan resolution and other scanning information
`o` Extracted image details (file type, pixel resolution, etc.)
- Information to be reported for offline procession of *en face* OCTA images for biomarker quantification

`o` Software utilized – ImageJ, Python, etc.
`o` Normalization method utilized (CLAHE or any other appropriate method)
`o` Binarization method applied (localized binarization methods preferred e.g., Wan, Bernsen, Wolf and Phansalkar)
- Biomarker quantification methods (not obtained from proprietary software):

`o` Detailed description and formula for biomarker estimation (such as VD, FD, etc.)
`o` Details of OCTA-ReVA, OCTAVA, or similar open-source tool usage (if applicable)
`o` Layer specific or custom biomarker details (if applicable)

## Conclusion

This study demonstrates that binarization algorithms substantially influence OCTA-derived vascular density metrics, with variability exceeding physiologic changes in some cases. Among the 29 algorithms evaluated Wan, Bernsen, Wolf and Phansalkar algorithms (local) were most consistent with Optovue benchmarks, while global methods introduced significant variability.

SCP layer was highly sensitive to binarization algorithms, whereas DCP layer exhibited greater resilience: a novel finding with implications for layer-specific analysis. The superior performance of local adaptive methods (normalization and binarization) underscores the need for stricter, standardized image processing protocols to ensure reproducibility across studies and devices. Future work should validate these findings in larger cohorts, include pathological eyes, and explore automated approaches for robust OCTA image binarization.

## Data Availability

The anonymized data collected are available via the University of Texas Southwestern Medical Center online data repository: https://dataverse.tdl.org/previewurl.xhtml?token=6c5114c7-f242-4d74-9791-8b35af395d0f Code repository: https://github.com/isi-11/OCTA-Binarization

## Acknowledgements

The authors would like to acknowledge the assistance of Angela Guillory and the staff members of Animal Resource Center at University of Texas Southwestern Medical Center, Dallas, TX. We extend our sincere gratitude to the clinical staff at the University of Texas Southwestern Medical Center for their invaluable assistance with data collection during this study.

## Author Contributions

Study conception or design – SSP, UH

Data acquisition – SSP, ISI, JSR, KA, SFE, MB, AD, AP, MD, JP, UH

Data analysis or interpretation – SSP, SI, JSR, SFE, UH

Manuscript writing – All

Funding – UH, JP

---

Table 1 (a). The average VD values for the SCP and DCP for both human and porcine subjects post-binarization with each algorithm. All VD values are reported as % mean ± standard deviation.

Table 1 (b). Pairwise comparison of Optovue VD values vs. binarization algorithm processed VD values for the SCP layer in human subjects (n=22).

Table 1 (c). Pairwise comparison of Optovue VD values vs. binarization algorithm processed VD values for the DCP layer in human subjects (n=22).

Table 1 (d). Pairwise comparison of Optovue VD values vs. binarization algorithm processed VD values for the SCP layer in porcine subjects (n=29).

Table 2. Comparative analysis of binarization techniques applied to OCTA scans in literature [12, 17, 76, 93–95].

