## Supplementary material for "Influence of Binarization Process on Vascular Density Metrics: A Quantitative Optical Coherence Tomography Angiography Assessment in Human and Porcine Retinas": Tables 1-2

| **Table 1 (a).** The VD values for the SCP and DCP layers for both human (n=22) and porcine (n=29) subjects, respectively, after binarization algorithm was applied. All VD values are reported as % (mean ± standard deviation). | | | | |
| --- | --- | --- | --- | --- |
| **Binarization Algorithm** | **SCP-Human (%)** | **DCP-Human (%)** | **SCP-Porcine (%)** | **DCP-Porcine (%)** |
| Original (Optovue) | 48.3 ± 1.4 | 51.9 ± 1.7 | 46.2 ± 1.8 | 50.0 ± 1.4 |
| Bataineh | 53.5 ± 1.2 | 54.4 ± 1.8 | 52.9 ± 1.4 | 51.5 ± 1.4 |
| Bernsen | 15.0 ± 2.0 | 19.0 ± 3.1 | 13.3 ± 1.8 | 15.3 ± 2.7 |
| ImageJ Default | 67.6 ± 4.2 | 56.2 ± 2.3 | 68.5 ± 5.1 | 61.2 ± 4.4 |
| Gatos | 51.0 ± 2.0 | 60.2 ± 2.6 | 50.4 ± 2.3 | 56.3 ± 2.2 |
| Huang | 48.5 ± 5.2 | 46.2 ± 3.5 | 46.9 ± 4.8 | 45.0 ± 3.4 |
| Intermodes | 50.5 ± 1.3 | 54.3 ± 1.3 | 50.3 ± 1.5 | 52.5 ± 1.1 |
| ISauvola | 43.0 ± 5.2 | 47.5 ± 3.1 | 44.5 ± 4.8 | 46.3 ± 5.4 |
| IsoData | 68.6 ± 4.1 | 56.1 ± 2.4 | 69.8 ± 4.5 | 59.0 ± 4.8 |
| Li | 55.6 ± 2.2 | 49.9 ± 2.1 | 55.3 ± 3.1 | 50.5 ± 2.4 |
| MaxEntropy | 91.1 ± 1.0 | 94.2 ± 1.6 | 91.4 ± 1.2 | 94.0 ± 0.7 |
| Mean | 56.3 ± 1.3 | 53.3 ± 1.9 | 56.7 ± 2.1 | 54.3 ± 1.6 |
| MinError | 43.2 ± 4.9 | 45.2 ± 4.8 | 42.0 ± 3.9 | 44.8 ± 4.6 |
| Minimum | 47.2 ± 8.1 | 46.9 ± 3.7 | 48.4 ± 5.9 | 47.0 ± 7.0 |
| Moments | 74.2 ± 2.2 | 68.2 ± 2.3 | 75.7 ± 2.1 | 70.9 ± 3.1 |
| Niblack | 34.7 ± 1.1 | 38.3 ± 0.9 | 34.7 ± 1.3 | 37.4 ± 1.3 |
| Nick | 57.0 ± 1.2 | 60.2 ± 1.3 | 56.2 ± 1.2 | 58.1 ± 1.2 |
| Otsu | 72.2 ± 2.7 | 59.4 ± 3.6 | 73.0 ± 3.8 | 62.3 ± 4.8 |
| Percentile | 50.4 ± 1.1 | 50.3 ± 1.1 | 50.3 ± 1.4 | 50.1 ± 1.5 |
| Phansalkar | 46.1 ± 1.8 | 50.7 ± 2.0 | 44.9 ± 2.3 | 47.2 ± 1.8 |
| Random Walker | 41.6 ± 2.8 | 54.2 ± 8.5 | 21.2 ± 3.8 | 46.4 ± 5.5 |
| Renyi Entropy | 90.7 ± 1.2 | 93.2 ± 2.1 | 91.3 ± 1.1 | 93.5 ± 0.7 |
| Sauvola | 50.5 ± 1.3 | 54.3 ± 1.3 | 49.6 ± 1.4 | 51.6 ± 1.0 |
| Shanbhag | 58.0 ± 1.2 | 56.0 ± 0.7 | 59.0 ± 1.1 | 54.8 ± 1.3 |
| Su | 90.2 ± 4.8 | 70.2 ± 2.9 | 91.8 ± 4.8 | 76.6 ± 6.8 |
| Triangle | 54.4 ± 1.4 | 57.8 ± 1.5 | 53.7 ± 1.5 | 55.8 ± 1.3 |
| TR Singh | 27.8 ± 5.6 | 35.0 ± 5.1 | 26.2 ± 3.7 | 19.3 ± 12.5 |
| Wan | 10.7 ± 1.0 | 11.3 ± 1.3 | 9.8 ± 0.9 | 10.0 ± 1.0 |
| Wolf | 46.5 ± 1.2 | 49.3 ± 0.9 | 46.3 ± 1.4 | 47.9 ± 0.8 |
| Yen | 91.4 ± 1.1 | 94.3 ± 1.2 | 91.8 ± 1.3 | 94.3 ± 0.8 |

**Table 1 (b).** Pairwise comparison of Optovue VD values vs. binarization algorithm processed VD values for the SCP layer in human subjects (n=22).

| **Comparison** | ***t-*value** | ***p-*value** | ***q*-value** | **Significant** |
| --- | --- | --- | --- | --- |
| Original vs. Bataineh | -18.2666 | 2.28E-14 | 4.50E-14 | TRUE |
| Original vs. Bernsen | 94.04263 | 4.66E-29 | 2.71E-28 | TRUE |
| Original vs. Default ImageJ | -18.2476 | 2.33E-14 | 4.50E-14 | TRUE |
| Original vs. Gatos | -6.46978 | 2.07E-06 | 3.00E-06 | TRUE |
| Original vs. Huang | -0.22319 | 0.825541 | 0.825541 | FALSE |
| Original vs. ISauvola | -6.33259 | 2.80E-06 | 3.53E-06 | TRUE |
| Original vs. Intermodes | 4.543935 | 0.000177 | 0.00019 | TRUE |
| Original vs. IsoData | -19.6715 | 5.22E-15 | 1.26E-14 | TRUE |
| Original vs. Li | -13.3656 | 9.73E-12 | 1.49E-11 | TRUE |
| Original vs. Max Entropy | -109.989 | 1.75E-30 | 2.25E-29 | TRUE |
| Original vs. Mean | -17.6007 | 4.75E-14 | 8.10E-14 | TRUE |
| Original vs. MinError | 4.865055 | 8.25E-05 | 9.20E-05 | TRUE |
| Original vs. Minimum | 0.602908 | 0.553026 | 0.572777 | FALSE |
| Original vs. Moments | -38.7477 | 5.07E-21 | 2.10E-20 | TRUE |
| Original vs. Niblack | 52.41373 | 9.49E-24 | 4.59E-23 | TRUE |
| Original vs. Nick | -27.0854 | 8.10E-18 | 2.35E-17 | TRUE |
| Original vs. Otsu | -32.5365 | 1.88E-19 | 6.05E-19 | TRUE |
| Original vs. Percentile | -5.58677 | 1.52E-05 | 1.84E-05 | TRUE |
| Original vs. Phansalkar | 6.416175 | 2.33E-06 | 3.21E-06 | TRUE |
| Original vs. Random Walker | 13.69416 | 6.15E-12 | 9.90E-12 | TRUE |
| Original vs. Renyi Entropy | -94.8869 | 3.87E-29 | 2.71E-28 | TRUE |
| Original vs. Sauvola | -6.33259 | 2.80E-06 | 3.53E-06 | TRUE |
| Original vs. Su | -23.1243 | 2.02E-16 | 5.33E-16 | TRUE |
| Original vs. Shanbhag | -35.5215 | 3.07E-20 | 1.11E-19 | TRUE |
| Original vs. TR Singh | -17.9847 | 3.10E-14 | 5.62E-14 | TRUE |
| Original vs. Triangle | 18.42498 | 1.92E-14 | 4.29E-14 | TRUE |
| Original vs. Wan | 140.9506 | 9.64E-33 | 2.79E-31 | TRUE |
| Original vs. Wolf | 5.277232 | 3.12E-05 | 3.62E-05 | TRUE |
| Original vs. Yen | -108.512 | 2.32E-30 | 2.25E-29 | TRUE |

**Table 1 (c).** Pairwise comparison of Optovue VD values vs. binarization algorithm processed VD values for the DCP layer in human subjects (n=22).

| **Comparison** | ***t-*value** | ***p-*value** | ***q*-value** | **Significant** |
| --- | --- | --- | --- | --- |
| Original vs. Bataineh | -8.3677 | 3.98E-08 | 7.21E-08 | TRUE |
| Original vs. Bernsen | 82.9096 | 6.53E-28 | 9.47E-27 | TRUE |
| Original vs. Default ImageJ | -6.33388 | 2.79E-06 | 3.86E-06 | TRUE |
| Original vs. Gatos | -25.1076 | 3.80E-17 | 1.38E-16 | TRUE |
| Original vs. Huang | 7.503877 | 2.26E-07 | 3.45E-07 | TRUE |
| Original vs. ISauvola | -8.91523 | 1.39E-08 | 2.69E-08 | TRUE |
| Original vs. Intermodes | 6.131731 | 4.39E-06 | 5.79E-06 | TRUE |
| Original vs. IsoData | -5.90076 | 7.40E-06 | 9.34E-06 | TRUE |
| Original vs. Li | 2.571026 | 0.017809 | 0.019128 | TRUE |
| Original vs. Max Entropy | -70.8017 | 1.78E-26 | 1.29E-25 | TRUE |
| Original vs. Mean | -2.46359 | 0.022474 | 0.023277 | TRUE |
| Original vs. MinError | 6.632146 | 1.45E-06 | 2.10E-06 | TRUE |
| Original vs. Minimum | 5.657899 | 1.29E-05 | 1.56E-05 | TRUE |
| Original vs. Moments | -21.1601 | 1.21E-15 | 3.51E-15 | TRUE |
| Original vs. Niblack | 45.64241 | 1.69E-22 | 8.18E-22 | TRUE |
| Original vs. Nick | -32.1551 | 2.39E-19 | 9.92E-19 | TRUE |
| Original vs. Otsu | -8.0552 | 7.37E-08 | 1.26E-07 | TRUE |
| Original vs. Percentile | 3.343747 | 0.003078 | 0.003434 | TRUE |
| Original vs. Phansalkar | 4.456523 | 0.000218 | 0.000253 | TRUE |
| Original vs. Random Walker | -1.53499 | 0.139716 | 0.139716 | FALSE |
| Original vs. Renyi Entropy | -57.7889 | 1.24E-24 | 7.18E-24 | TRUE |
| Original vs. Sauvola | -8.91523 | 1.39E-08 | 2.69E-08 | TRUE |
| Original vs. Su | -10.7306 | 5.56E-10 | 1.24E-09 | TRUE |
| Original vs. Shanbhag | -20.1573 | 3.21E-15 | 8.45E-15 | TRUE |
| Original vs. TR Singh | -21.9565 | 5.76E-16 | 1.85E-15 | TRUE |
| Original vs. Triangle | 18.32705 | 2.14E-14 | 5.16E-14 | TRUE |
| Original vs. Wan | 213.5196 | 1.58E-36 | 4.58E-35 | TRUE |
| Original vs. Wolf | 7.796885 | 1.24E-07 | 2.00E-07 | TRUE |
| Original vs. Yen | -79.3572 | 1.63E-27 | 1.58E-26 | TRUE |

**Table 1 (d).** Pairwise comparison of Optovue VD values vs. binarization algorithm processed VD values for the SCP layer in porcine subjects (n=29).

| **Comparison** | ***t-*value** | ***p-*value** | ***q*-value** | **Significant** |
| --- | --- | --- | --- | --- |
| Original vs. Bataineh | -18.8913 | 1.80E-17 | 3.07E-17 | TRUE |
| Original vs. Bernsen | 134.8757 | 6.13E-41 | 4.44E-40 | TRUE |
| Original vs. Default ImageJ | -18.9983 | 1.55E-17 | 2.82E-17 | TRUE |
| Original vs. Gatos | -13.2737 | 1.33E-13 | 2.03E-13 | TRUE |
| Original vs. Huang | -0.73924 | 0.465916 | 0.482556 | FALSE |
| Original vs. ISauvola | -13.1895 | 1.55E-13 | 2.25E-13 | TRUE |
| Original vs. Intermodes | 1.756101 | 0.090006 | 0.096673 | FALSE |
| Original vs. IsoData | -21.2551 | 8.17E-19 | 1.58E-18 | TRUE |
| Original vs. Li | -11.9909 | 1.52E-12 | 2.09E-12 | TRUE |
| Original vs. Max Entropy | -133.385 | 8.36E-41 | 4.85E-40 | TRUE |
| Original vs. Mean | -17.1932 | 2.05E-16 | 3.31E-16 | TRUE |
| Original vs. MinError | 5.305067 | 1.21E-05 | 1.46E-05 | TRUE |
| Original vs. Minimum | -1.8849 | 0.069861 | 0.077922 | FALSE |
| Original vs. Moments | -43.6715 | 2.65E-27 | 9.61E-27 | TRUE |
| Original vs. Niblack | 53.55362 | 9.36E-30 | 4.53E-29 | TRUE |
| Original vs. Nick | -37.2889 | 2.06E-25 | 5.97E-25 | TRUE |
| Original vs. Otsu | -27.8286 | 6.03E-22 | 1.34E-21 | TRUE |
| Original vs. Percentile | -10.8263 | 1.62E-11 | 2.04E-11 | TRUE |
| Original vs. Phansalkar | 4.284434 | 0.000195 | 0.000226 | TRUE |
| Original vs. Random Walker | 48.04144 | 1.90E-28 | 7.87E-28 | TRUE |
| Original vs. Renyi Entropy | -136.473 | 4.41E-41 | 4.26E-40 | TRUE |
| Original vs. Sauvola | -11.4097 | 4.85E-12 | 6.39E-12 | TRUE |
| Original vs. Su | -36.3684 | 4.09E-25 | 1.08E-24 | TRUE |
| Original vs. Shanbhag | -40.2579 | 2.50E-26 | 8.06E-26 | TRUE |
| Original vs. TR Singh | -27.4089 | 9.09E-22 | 1.88E-21 | TRUE |
| Original vs. Triangle | 28.65824 | 2.72E-22 | 6.57E-22 | TRUE |
| Original vs. Wan | 141.238 | 1.69E-41 | 2.45E-40 | TRUE |
| Original vs. Wolf | -0.33734 | 0.738379 | 0.738379 | FALSE |
| Original vs. Yen | -143.299 | 1.13E-41 | 2.45E-40 | TRUE |

**Table 1 (e).** Pairwise comparison of Optovue VD values vs. binarization algorithm processed VD values for the DCP layer in porcine subjects (n=29).

| **Comparison** | ***t-*value** | ***p-*value** | ***q*-value** | **Significant** |
| --- | --- | --- | --- | --- |
| Original vs. Bataineh | -7.062 | 1.11E-07 | 1.46E-07 | TRUE |
| Original vs. Bernsen | 73.83587 | 1.24E-33 | 7.19E-33 | TRUE |
| Original vs. Default ImageJ | -12.5461 | 5.18E-13 | 1.00E-12 | TRUE |
| Original vs. Gatos | -15.4635 | 3.04E-15 | 7.35E-15 | TRUE |
| Original vs. Huang | 8.863862 | 1.29E-09 | 1.86E-09 | TRUE |
| Original vs. ISauvola | -10.082 | 8.03E-11 | 1.37E-10 | TRUE |
| Original vs. Intermodes | 3.563889 | 0.001335 | 0.001489 | TRUE |
| Original vs. IsoData | -9.7763 | 1.58E-10 | 2.41E-10 | TRUE |
| Original vs. Li | -1.21615 | 0.234081 | 0.242441 | FALSE |
| Original vs. Max Entropy | -168.861 | 1.14E-43 | 2.44E-42 | TRUE |
| Original vs. Mean | -10.0937 | 7.83E-11 | 1.37E-10 | TRUE |
| Original vs. MinError | 6.690625 | 2.92E-07 | 3.68E-07 | TRUE |
| Original vs. Minimum | 2.225611 | 0.034268 | 0.036807 | TRUE |
| Original vs. Moments | -30.1903 | 6.62E-23 | 2.40E-22 | TRUE |
| Original vs. Niblack | 38.99116 | 6.03E-26 | 2.91E-25 | TRUE |
| Original vs. Nick | -37.672 | 1.55E-25 | 6.44E-25 | TRUE |
| Original vs. Otsu | -13.1771 | 1.59E-13 | 3.29E-13 | TRUE |
| Original vs. Percentile | -0.3243 | 0.748125 | 0.748125 | FALSE |
| Original vs. Phansalkar | 9.800725 | 1.50E-10 | 2.41E-10 | TRUE |
| Original vs. Random Walker | 3.673001 | 0.001002 | 0.001163 | TRUE |
| Original vs. Renyi Entropy | -156.765 | 9.14E-43 | 8.84E-42 | TRUE |
| Original vs. Sauvola | -6.64764 | 3.26E-07 | 3.94E-07 | TRUE |
| Original vs. Su | -25.3254 | 7.66E-21 | 2.22E-20 | TRUE |
| Original vs. Shanbhag | -19.7691 | 5.50E-18 | 1.45E-17 | TRUE |
| Original vs. TR Singh | -26.1372 | 3.28E-21 | 1.06E-20 | TRUE |
| Original vs. Triangle | 14.17678 | 2.65E-14 | 5.92E-14 | TRUE |
| Original vs. Wan | 146.8731 | 5.66E-42 | 4.10E-41 | TRUE |
| Original vs. Wolf | 7.401385 | 4.64E-08 | 6.40E-08 | TRUE |
| Original vs. Yen | -166.535 | 1.69E-43 | 2.44E-42 | TRUE |

**Table 2.** Comparative analysis of binarization techniques applied to OCTA scans in literature [12, 17, 76, 93-95].

| **Reference** | **Subject(s)** | **OCTA Images** | **Binarization techniques** | **Metrics** | **Main findings** |
| --- | --- | --- | --- | --- | --- |
| Our study | Human, Porcine | Superficial capillary plexus (SCP), deep capillary plexus (DCP) | 29 algorithms evaluated (See Fig. 3(a)-(d)) | Vascular density (VD) | Local, adaptive algorithms (Wan, Bernsen, Wolf and Phansalkar) outperform global ones; Percentile algorithm being exceptional for porcine DCP layer  No single technique fits all retinal layers or models universally |
| Corvi et al. 2020 | Human | SCP | 5 algorithms utilized - Default, Huang, IsoData, Mean, Otsu | VD | Thresholding algorithms had a significant effect on mean measurements (p < 0.0001), with all pairwise comparisons differing except the IsoData–Otsu comparison (p > 0.270).  Brightness perturbations of ±50% likewise yielded statistically significant differences (p < 0.0001). |
| Borrelli et al. 2021 | Human | SCP, deep vascular complex (DVC) | 6 algorithms utilized - global Default, global Mean, global Otsu, local mean, local Phansalkar, complex/multistep | Perfusion Density (PD), Vessel length density (VLD), Interclass Correlation Coefficient (ICC) | Global thresholding yielded better-quality binarized images than local or multistep methods, and the median filter most often provided the best histogram adjustment.  Local thresholds produced higher OCTA metrics, with differences between global and local methods influenced by diabetic macular edema and signal strength index.  Thresholding algorithm choice also significantly affected OCTA metric differences between healthy and diabetic retinopathy eyes. |
| Terheyden et al. 2020 | Human | SCP, DCP | 7 algorithms utilized - manual thresholding, Huang, Li, Otsu, Moments, Mean, Percentile | VD, Vessel Skeleton Density (VSD), ICC | Automated thresholding algorithms improve reproducibility of OCTA parameters and enhance sensitivity for detecting macular pathology, but different algorithms are not interchangeable.  The Mean algorithm warrants further investigation, and although automated methods are preferable, greater standardization is required for clinical application. |
| Arrigo et al. 2021 | Human | SCP, DCP, Choriocapillaris (CC) | 13 algorithms utilized - Default, Huang, Intermodes, Li, Max Entropy, Mean, Moments, Otsu, Percentile, Renyi Entropy, Shanbhag, Yen, and the fixed threshold established by the first experiment | Overlapping percentages between binarized and original images, VD, vessel tortuosity (VT), vessel dispersion (V_disp_), Foveal Avascular Zone (FAZ) detection | Huang, Li, Mean and Percentile were the most reliable binarization thresholds (p < 0.05), whereas the worst binarization thresholds were Intermodes, MaxEntropy, RenyiEntropy and Yen (p < 0.05).  All the thresholds variably underestimated VD metric and FAZ detection, with respect to the original OCTA images, whereas VT and V_disp_ metrics turned out to be more stable. |
| Freedman et al. 2022 | Human | SCP | 4 algorithms utilized: global Huan, global Otsu, local Niblack, local Phansalkar  Performed Background Subtraction, FAZ adjustment, and/or CLAHE, prior to binarization | VD, skeletonized vessel density (SVD),  fractal dimensions (FD) | Image processing methods can influence conclusions drawn from quantitative OCTA analyses, so results should be interpreted with caution.  Background subtraction or CLAHE may help support more standardized image processing. |
| Angeli et al. 2023 | Human | SCP | 2 algorithms utilized - Mexican Hat Filter, Shanbhag | VD, Skeleton Density, Vessel Diameter Index | Both binarization methods (Mexican Hat filer and Shanbhag) can segment the vascular network and quantify VD, but they yield substantially inconsistent results.  Determining the clinical relevance of these differences requires careful understanding of the image analysis approach.  For longitudinal monitoring, using the same processing method is recommended. |
| Cheng et al. 2023 | Human | Full retinal layer | Default + Mexican Hat Filter (enhances edges) | VD, VT, FD | Use of a Mexican hat filter was required to quantify capillary microvascular parameters, and excluding the FAZ was essential for evaluating VT. |
| Prangel et al. 2023 | Human | SCP, DCP, CC | 5 algorithms utilized - Default (ImageJ), Huang, IsoData, Mean, Otsu | VD | Automated thresholding algorithms cannot be interchanged.  Highlights the need for layer specific algorithm, especially CC layer. |
