## Supplemental Tables S1-S4 for "Influence of Binarization Process on Vascular Density Metrics: A Quantitative Optical Coherence Tomography Angiography Assessment in Human and Porcine Retinas"

Supplementary information for the manuscript

### Supplementary information:

### S1. CLAHE Table

To optimize Contrast Limited Adaptive Histogram Equalization (CLAHE) for our OCTA scans, we conducted a systematic analysis of block size and histogram bin size combinations, using peak signal-to-noise ratio (PSNR) as the primary outcome measure. PSNR was selected because it quantifies the fidelity of a processed image relative to the original, with higher values indicating less distortion and better image quality. The CLAHE slope parameter (clip limit in skimage.exposure.equalize_adapthist) did not meaningfully affect PSNR or image appearance in preliminary testing; therefore, it was retained at the default value of 0.01. We first varied block size while holding the histogram bin size constant at its default value of 256. Applying each parameter set across all OCTA images revealed that block sizes in the range of approximately 100 to 120 pixels produced the highest PSNR values (Table S1(a)). Next, we varied the histogram bin size while keeping the block size at the default value (none). This analysis demonstrated that a bin size of 100 consistently yielded the highest PSNR (Table S1(b)). Finally, we evaluated combined effects by fixing the block size at 100, 110, and 120 pixels while varying the bin size during CLAHE normalization. When the bin size was set to 100, block size had only a minimal influence on PSNR, with differences between the tested block sizes falling within a narrow range (Table S1(c)). However, a block size of 110 pixels was selected as the final parameter because it provided the most stable and reproducible PSNR across images, with slightly reduced variability compared with 100 or 120 pixels and produced visually uniform contrast enhancement without over-amplification of local noise. Thus, the final parameters for CLAHE normalization were set with a block size of 110 pixels, a histogram bin size of 100, and a slope of 0.01. The following tables show randomized, exemplary images utilized for optimizing the parameters for CLAHE normalization.

**Table S1(a).** The PSNR values for the SCP and DCP for both human and porcine subjects during CLAHE optimization while varying block size and holding bin size constant at its default value (256). All PSNR values are reported as dB.

| Sample Image No. | Block Size | SCP-Human | DCP-Human | SCP-Porcine | DCP-Porcine |
|---|---|---|---|---|---|
| 1 | 127 | 38.1972 | 31.566 | 38.4241 | 41.1269 |
| 1 | 120 | 38.3205 | 32.3875 | 38.5484 | 41.1471 |
| 1 | 110 | 38.3398 | 33.0993 | 38.6709 | 41.0223 |
| 1 | 100 | 38.4263 | 32.9718 | 38.5376 | 41.2749 |
| 1 | 90 | 38.0928 | 33.3892 | 38.3174 | 41.1887 |
| 1 | 80 | 38.0771 | 33.2419 | 38.1892 | 40.9268 |
| 1 | 70 | 37.5746 | 33.3338 | 37.8901 | 40.4736 |
| 1 | 60 | 37.4126 | 33.8108 | 37.2313 | 40.0445 |
| 1 | 50 | 36.4266 | 34.1776 | 36.5805 | 39.069 |
| 1 | 40 | 35.3996 | 33.1537 | 35.2879 | 35.1532 |
| 1 | 30 | 31.6402 | 35.057 | 31.4576 | 30.0515 |
| 1 | 20 | 33.7677 | 34.9112 | 34.945 | 38.1164 |
| 1 | 10 | 22.7732 | 25.5241 | 28.1117 | 31.7905 |
| 2 | 127 | 38.2097 | 41.0675 | 38.5153 | 39.0198 |
| 2 | 120 | 38.1971 | 41.1955 | 38.6127 | 38.2838 |
| 2 | 110 | 38.378 | 41.1914 | 38.6174 | 38.4954 |
| 2 | 100 | 38.3446 | 41.3528 | 38.3772 | 38.578 |
| 2 | 90 | 38.3257 | 41.0427 | 38.0805 | 39.5706 |
| 2 | 80 | 38.0131 | 40.501 | 38.1165 | 38.3713 |
| 2 | 70 | 37.7033 | 40.953 | 38.0501 | 38.1841 |
| 2 | 60 | 37.5192 | 39.6755 | 37.0714 | 37.9111 |
| 2 | 50 | 36.266 | 40.1577 | 36.8173 | 37.6376 |

| | | | | | |
|---|---|---|---|---|---|
| 2 | 40 | 34.9432 | 34.3106 | 34.2832 | 34.7601 |
| 2 | 30 | 31.2596 | 29.6593 | 30.5005 | 30.3422 |
| 2 | 20 | 35.2592 | 37.3746 | 35.3963 | 36.9907 |
| 2 | 10 | 28.6205 | 27.3552 | 28.274 | 27.8836 |
| 3 | 127 | 37.5369 | 37.1772 | 38.3542 | 40.3569 |
| 3 | 120 | 37.6309 | 37.4051 | 38.3627 | 40.1519 |
| 3 | 110 | 37.7142 | 37.2629 | 38.5148 | 40.0745 |
| 3 | 100 | 37.7372 | 37.2339 | 38.2209 | 40.2203 |
| 3 | 90 | 37.4941 | 37.3784 | 37.9999 | 39.7385 |
| 3 | 80 | 37.4107 | 37.1684 | 37.7991 | 39.5469 |
| 3 | 70 | 37.1915 | 37.0341 | 37.7479 | 39.7331 |
| 3 | 60 | 36.5221 | 36.498 | 36.8331 | 38.8153 |
| 3 | 50 | 35.9209 | 36.2376 | 36.5254 | 38.762 |
| 3 | 40 | 34.352 | 33.084 | 34.0097 | 35.0309 |
| 3 | 30 | 31.1525 | 29.244 | 30.7102 | 30.1153 |
| 3 | 20 | 35.0589 | 34.7348 | 34.5836 | 36.8661 |
| 3 | 10 | 27.1954 | 27.4017 | 28.5649 | 28.1934 |

**Table S1(b).** The PSNR values for the SCP and DCP for both human and porcine subjects during CLAHE optimization while varying bin size and holding block size constant at its default value (none). All PSNR values are reported as dB.

| Sample Image No. | Bin Size | SCP-Human | DCP-Human | SCP-Porcine | DCP-Porcine |
|---|---|---|---|---|---|
| 1 | 256 | 38.3398 | 33.0993 | 38.6709 | 41.0223 |
| 1 | 200 | 38.4789 | 35.648 | 39.1467 | 41.3446 |
| 1 | 150 | 35.449 | 38.4695 | 35.836 | 35.637 |
| 1 | 100 | 47.3939 | 47.496 | 47.5117 | 46.5865 |
| 1 | 75 | 45.4739 | 44.9986 | 43.2567 | 45.3383 |
| 1 | 50 | 41.6534 | 42.0077 | 41.6251 | 40.7769 |
| 1 | 25 | 35.5565 | 36.1523 | 34.8752 | 35.6396 |
| 1 | 15 | 31.6038 | 30.4175 | 30.5684 | 31.0293 |
| 1 | 10 | 27.224 | 27.6734 | 27.9695 | 27.6325 |
| 2 | 256 | 38.378 | 41.1914 | 38.6174 | 38.4954 |
| 2 | 200 | 38.7555 | 41.2079 | 39.2558 | 39.7787 |
| 2 | 150 | 34.2032 | 36.6983 | 34.9511 | 37.1534 |
| 2 | 100 | 47.3095 | 49.1804 | 48.0589 | 48.1539 |
| 2 | 75 | 43.1931 | 46.7482 | 47.7307 | 49.5732 |
| 2 | 50 | 41.5696 | 41.6973 | 42.144 | 41.8904 |
| 2 | 25 | 34.8401 | 35.8629 | 36.6978 | 37.0507 |
| 2 | 15 | 30.5863 | 31.2115 | 30.2947 | 32.049 |
| 2 | 10 | 27.9794 | 27.6639 | 27.4868 | 27.6728 |
| 3 | 256 | 37.7142 | 37.2629 | 38.5148 | 40.0745 |
| 3 | 200 | 38.1633 | 38.4447 | 38.6762 | 40.3379 |
| 3 | 150 | 34.48 | 36.4473 | 35.4214 | 36.9566 |
| 3 | 100 | 46.7849 | 48.2705 | 47.1039 | 48.1735 |
| 3 | 75 | 43.3838 | 45.1453 | 42.2961 | 48.9667 |
| 3 | 50 | 40.8059 | 42.7879 | 41.2186 | 41.878 |
| 3 | 25 | 34.8078 | 36.974 | 34.9068 | 36.865 |
| 3 | 15 | 29.7539 | 29.9089 | 30.4401 | 32.0106 |
| 3 | 10 | 27.5446 | 27.5083 | 27.8943 | 27.6435 |

**Table S1(c).** The PSNR values for the SCP and DCP for both human and porcine subjects during CLAHE optimization while varying bin size and holding block size constant at 110 pixels. All PSNR values are reported as dB.

| Sample Image No. | Block Size | Bin Size | SCP-Human | DCP-Human | SCP-Porcine | DCP-Porcine |
|---|---|---|---|---|---|---|
| 1 | 110 | 256 | 38.3398 | 33.0993 | 38.6709 | 41.0223 |
| 1 | 110 | 200 | 38.4789 | 35.648 | 39.1467 | 41.3446 |
| 1 | 110 | 150 | 35.449 | 38.4695 | 35.836 | 35.637 |
| 1 | 110 | 100 | 47.3939 | 47.496 | 47.5117 | 46.5865 |
| 1 | 110 | 75 | 45.4739 | 44.9986 | 43.2567 | 45.3383 |
| 1 | 110 | 50 | 41.6534 | 42.0077 | 41.6251 | 40.7769 |
| 1 | 110 | 25 | 35.5565 | 36.1523 | 34.8752 | 35.6396 |
| 1 | 110 | 15 | 31.6038 | 30.4175 | 30.5684 | 31.0293 |
| 1 | 110 | 10 | 27.224 | 27.6734 | 27.9695 | 27.6325 |
| 2 | 110 | 256 | 38.378 | 41.1914 | 38.6174 | 38.4954 |
| 2 | 110 | 200 | 38.7555 | 41.2079 | 39.2558 | 39.7787 |
| 2 | 110 | 150 | 34.2032 | 36.6983 | 34.9511 | 37.1534 |
| 2 | 110 | 100 | 47.3095 | 49.1804 | 48.0589 | 48.1539 |
| 2 | 110 | 75 | 43.1931 | 46.7482 | 47.7307 | 49.5732 |
| 2 | 110 | 50 | 41.5696 | 41.6973 | 42.144 | 41.8904 |
| 2 | 110 | 25 | 34.8401 | 35.8629 | 36.6978 | 37.0507 |
| 2 | 110 | 15 | 30.5863 | 31.2115 | 30.2947 | 32.049 |
| 2 | 110 | 10 | 27.9794 | 27.6639 | 27.4868 | 27.6728 |
| 3 | 110 | 256 | 37.7142 | 37.2629 | 38.5148 | 40.0745 |
| 3 | 110 | 200 | 38.1633 | 38.4447 | 38.6762 | 40.3379 |
| 3 | 110 | 150 | 34.48 | 36.4473 | 35.4214 | 36.9566 |
| 3 | 110 | 100 | 46.7849 | 48.2705 | 47.1039 | 48.1735 |
| 3 | 110 | 75 | 43.3838 | 45.1453 | 42.2961 | 48.9667 |
| 3 | 110 | 50 | 40.8059 | 42.7879 | 41.2186 | 41.878 |
| 3 | 110 | 25 | 34.8078 | 36.974 | 34.9068 | 36.865 |
| 3 | 110 | 15 | 29.7539 | 29.9089 | 30.4401 | 32.0106 |
| 3 | 110 | 10 | 27.5446 | 27.5083 | 27.8943 | 27.6435 |

**S2. Pixel Signal-to-Noise ratio (PSNR) quantified for each binarization algorithm.**

**Table S2.** The PSNR values for the SCP and DCP for both human and porcine subjects post-binarization with each algorithm. All PSNR values are reported as dB (mean ± standard deviation).

| Binarization Algorithm | SCP-Human | DCP-Human | SCP-Porcine | DCP-Porcine |
|---|---|---|---|---|
| Bataineh | 7.0 ± 0.2 | 7.4 ± 0.2 | 6.7 ± 0.3 | 7.2 ± 0.3 |
| Bernsen | 11.1 ± 0.3 | 10.3 ± 0.3 | 11.4 ± 0.4 | 10.8 ± 0.4 |
| ImageJ Default | 2.5 ± 0.2 | 3.0 ± 0.1 | 2.4 ± 0.2 | 2.7 ± 0.2 |
| Gatos | 7.1 ± 0.2 | 6.5 ± 0.1 | 6.9 ± 0.2 | 6.5 ± 0.1 |
| Huang | 3.2 ± 0.3 | 3.3 ± 0.2 | 3.2 ± 0.3 | 3.3 ± 0.2 |
| Intermodes | 7.5 ± 0.2 | 7.5 ± 0.2 | 7.3 ± 0.2 | 7.3 ± 0.2 |
| ISauvola | 3.4 ± 0.3 | 3.3 ± 0.1 | 3.4 ± 0.3 | 3.2 ± 0.3 |
| IsoData | 2.5 ± 0.2 | 3.0 ± 0.1 | 2.4 ± 0.2 | 2.7 ± 0.2 |
| Li | 2.9 ± 0.1 | 3.2 ± 0.1 | 2.8 ± 0.2 | 3.0 ± 0.1 |
| MaxEntropy | 2.3 ± 0.2 | 2.8 ± 0.2 | 2.2 ± 0.2 | 2.4 ± 0.2 |
| Mean | 2.8 ± 0.1 | 3.1 ± 0.1 | 2.8 ± 0.1 | 2.9 ± 0.1 |
| MinError | 3.4 ± 0.2 | 3.4 ± 0.2 | 3.5 ± 0.2 | 3.3 ± 0.2 |
| Minimum | 3.2 ± 0.5 | 3.3 ± 0.2 | 3.2 ± 0.4 | 3.2 ± 0.4 |
| Moments | 2.4 ± 0.1 | 2.7 ± 0.2 | 2.3 ± 0.1 | 2.5 ± 0.1 |
| Niblack | 9.4 ± 0.1 | 9.1 ± 0.1 | 9.2 ± 0.1 | 9.1 ± 0.1 |
| Nick | 6.7 ± 0.2 | 7.0 ± 0.2 | 6.6 ± 0.2 | 6.7 ± 0.2 |
| Otsu | 2.4 ± 0.1 | 2.9 ± 0.2 | 2.3 ± 0.2 | 2.7 ± 0.2 |
| Percentile | 3.1 ± 0.1 | 3.2 ± 0.1 | 3.1 ± 0.1 | 3.1 ± 0.1 |
| Phansalkar | 8.0 ± 0.2 | 7.9 ± 0.1 | 7.9 ± 0.2 | 7.9 ± 0.2 |
| Random Walker | 10.0 ± 0.6 | 8.0 ± 0.9 | 10.1 ± 0.6 | 9.0 ± 0.7 |
| Renyi Entropy | 2.3 ± 0.2 | 2.8 ± 0.2 | 2.2 ± 0.2 | 2.4 ± 0.2 |
| Sauvola | 7.5 ± 0.2 | 7.5 ± 0.2 | 7.4 ± 0.2 | 7.5 ± 0.2 |
| Shanbhag | 6.5 ± 0.3 | 7.2 ± 0.3 | 6.1 ± 0.3 | 6.9 ± 0.3 |
| Su | 2.3 ± 0.1 | 2.7 ± 0.2 | 2.2 ± 0.1 | 2.4 ± 0.2 |
| Triangle | 7.0 ± 0.2 | 7.2 ± 0.2 | 6.9 ± 0.2 | 7.0 ± 0.2 |
| TR Singh | 4.5 ± 0.5 | 3.9 ± 0.4 | 4.7 ± 0.3 | 5.5 ± 1.4 |
| Wan | 11.0 ± 0.4 | 10.0 ± 0.4 | 11.3 ± 0.5 | 10.7 ± 0.4 |
| Wolf | 8.0 ± 0.2 | 8.1 ± 0.2 | 7.8 ± 0.3 | 7.9 ± 0.2 |
| Yen | 2.3 ± 0.2 | 2.8 ± 0.2 | 2.2 ± 0.2 | 2.4 ± 0.2 |

### S3. Performance (or deviation) analysis of VD values from 29 binarization algorithms

We showed the comparison of mean VD values - ground truth, in comparison with all the calculated VDs from respective algorithms. A performance graph (or the magnitude of deviation) for each binarization algorithm is shown here by plotting the root mean square error (RMSE) for each algorithm vs. ground truth VD values in each respective layer of the given cohort

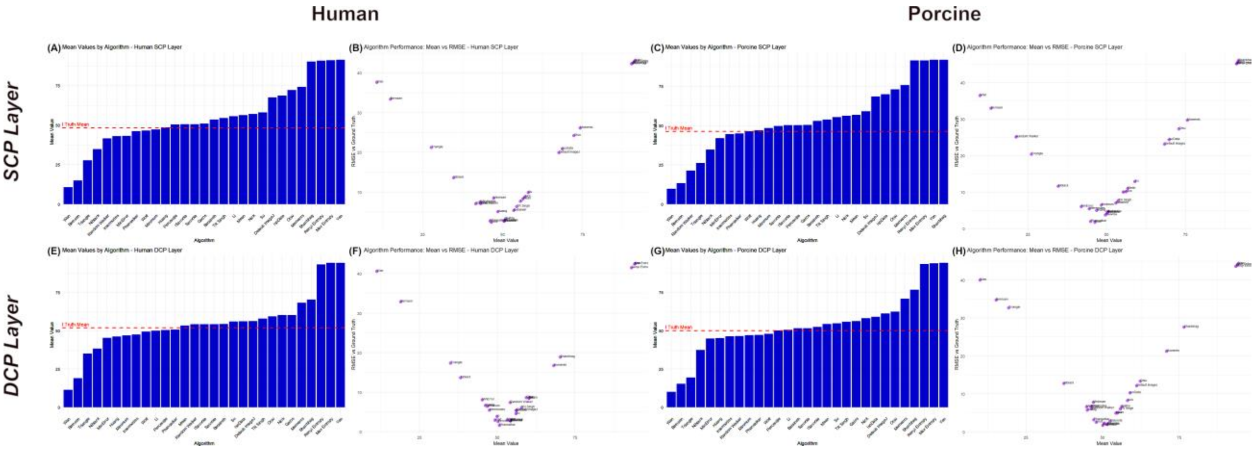

**Figure S3.** Mean vascular density (VD) and root mean square error (RMSE) from Optovue (Ground Truth) compared to other binarization algorithms. Mean VD values are compared across the algorithms for the SCP (human SCP (A), porcine (C)) and DCP (human DCP (E), porcine DCP (G)) layers with the Ground Truth or Optovue mean displayed in a dashed red line. Further, we plot the VD mean vs. RMSE values for both the retinal layers sourced from human (B & F) and porcine (D & H) subjects, respectively.

**S4. Interclass correlation (ICC) analysis – Optovue vs. binarization algorithms.**

Inter-algorithm reliability was evaluated using interclass correlation analysis (ICC) methodology. Vascular density measurements (human (n=22) and porcine scans (n=29)) obtained from Optovue machine (source) were compared to the vascular density values obtained after post-processing the same scans with 29 binarization methods (see *Materials and Methods* for details) using robust ICC method - *pysch* package in *R Studio* environment. Agreements between Optovue measures and each binarization method were quantified using means of ICC values. ICC values of 0.9 or greater were considered *Strong*, between 0.75-0.9 were considered *Excellent,* between 0.6-0.75 were considered *Good*, between 0.4-0.6 were considered *Fair*, and below 0.4 were considered *Poor*.

a. ICC analysis - Human superficial capillary plexus (SCP).

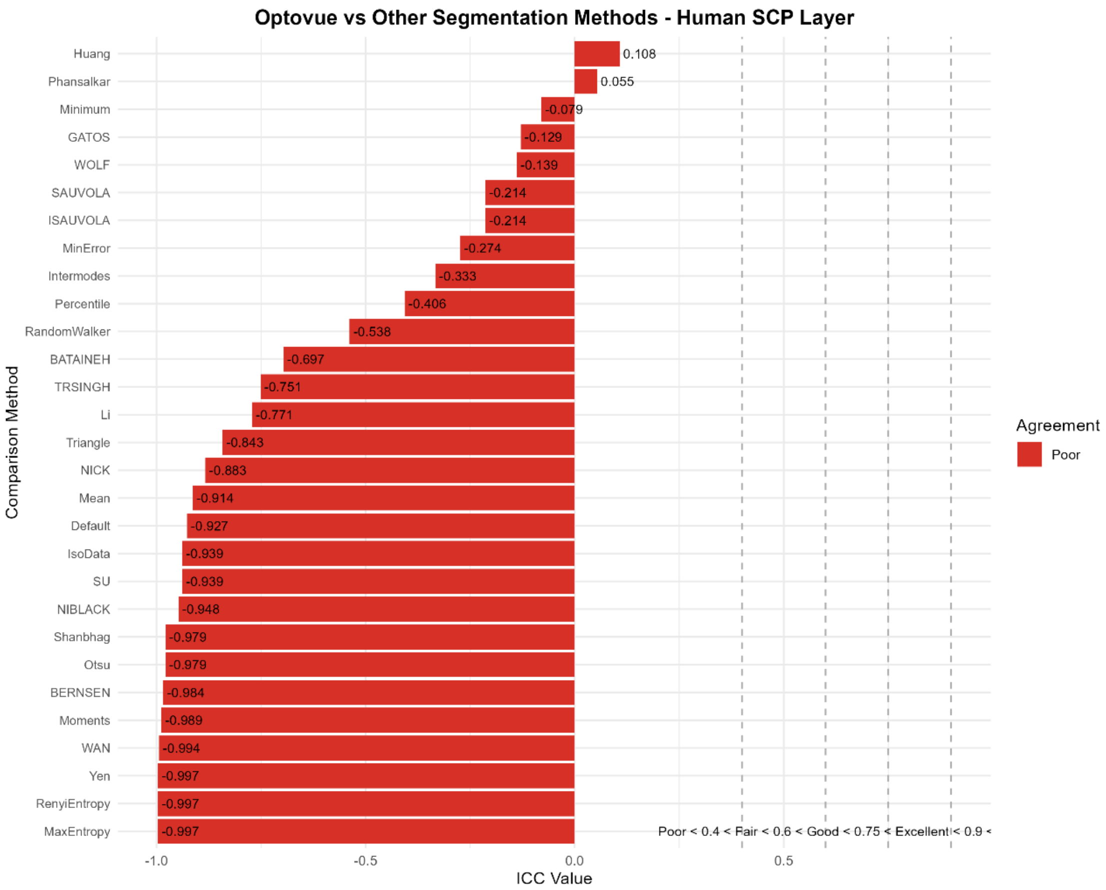

Fig.S4(a) Comparison of Optovue with other binarization methods for human superficial capillary plexus (SCP) is shown here. Huang (ρ = 0.108) and Phansalkar (ρ = 0.055) methods had poor agreements, whereas the rest fared worse when compared Optovue obtain vascular density values for human SCP scans.

b. ICC analysis - Human deep capillary plexus (DCP).

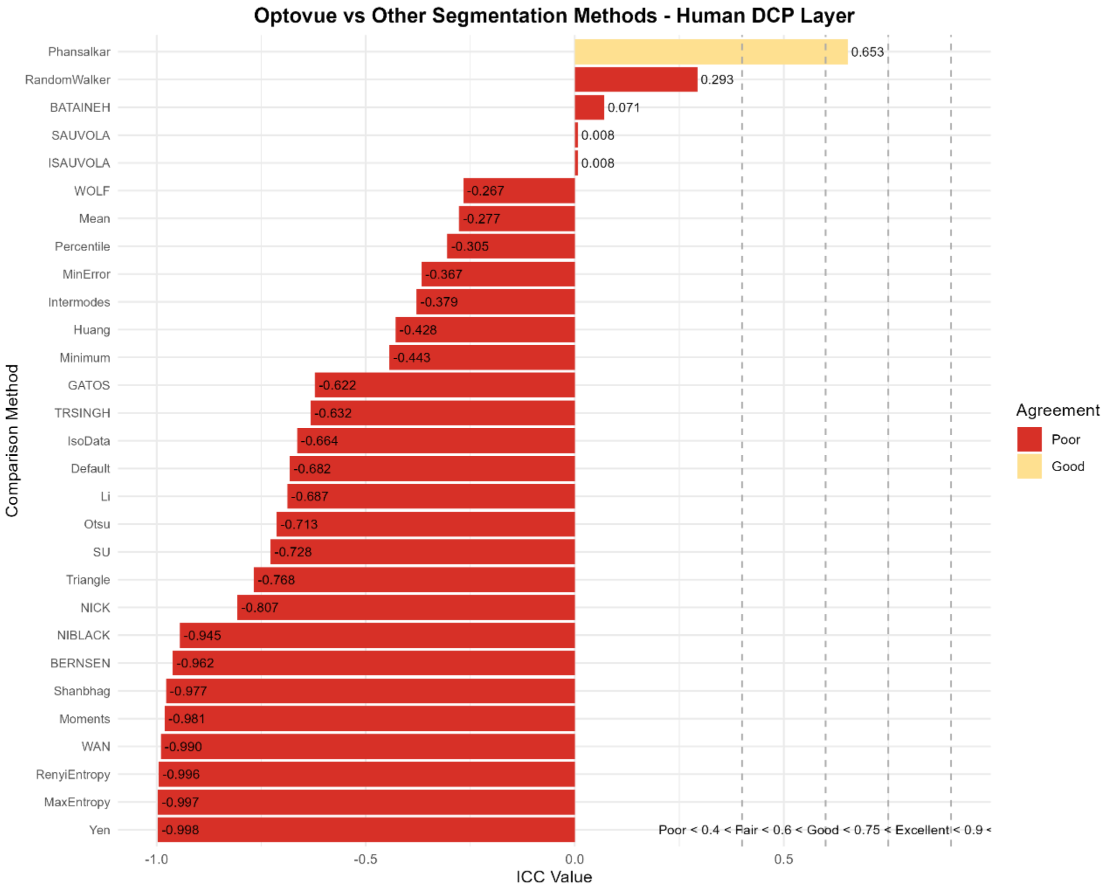

Fig.S4(b) Comparison of Optovue with other binarization methods for human deep capillary plexus (DCP) is shown here. Phansalkar (ρ = 0.653) had the best correlation with Optovue vascular density measurements. RandomWalker (ρ = 0.293) and Bataineh (ρ = 0.293) methods had poor agreements, where as the rest of the binarization methods fared worse when compared Optovue obtain vascular density values for human SCP scans.

c. ICC analysis - Porcine superficial capillary plexus (SCP).

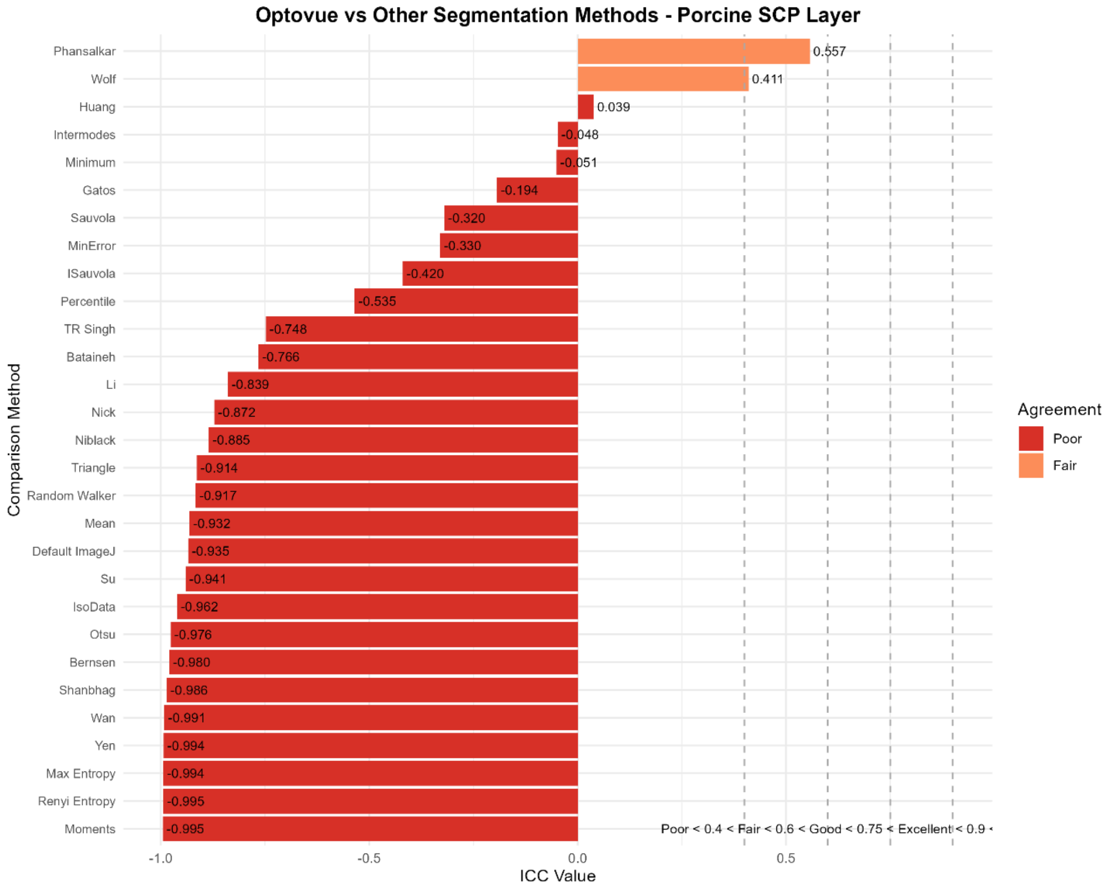

Fig.S4(c) Comparison of Optovue with other binarization methods for porcine superficial capillary plexus (SCP) is shown here. Phansalkar ($\rho = 0.557$) had a *Fair* correlation with Optovue vascular density measurements. Along the same lines, Wolf ($\rho = 0.411$) had fairly similar ICC values. The rest of the binarization procedures did not provide a good ICC agreement with Optovue values.

d. ICC analysis - Porcine deep capillary plexus (DCP).

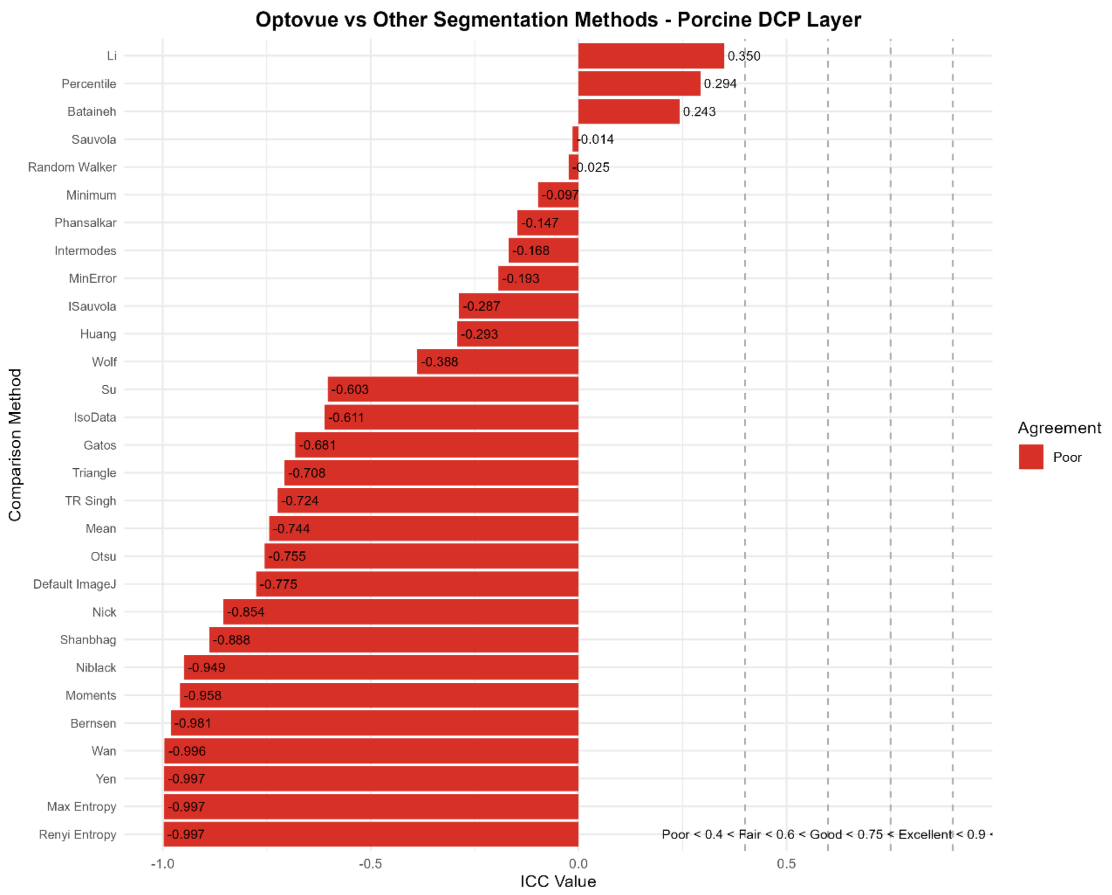

Fig. S4(d) Comparison of Optovue with other binarization methods for porcine deep capillary plexus (SCP) is shown here. Comparatively, none of the binarization methods exhibited any agreement with Optovue values.
